# Fifty Ways to Tag your Pubtypes: Multi-Tagger, a Set of Probabilistic Publication Type and Study Design Taggers to Support Biomedical Indexing and Evidence-Based Medicine

**DOI:** 10.1101/2021.07.13.21260468

**Authors:** Aaron M. Cohen, Jodi Schneider, Yuanxi Fu, Marian S. McDonagh, Prerna Das, Arthur W. Holt, Neil R. Smalheiser

## Abstract

**Objective:** Indexing articles according to publication types (PTs) and study designs can be a great aid to filtering literature for information retrieval, especially for evidence syntheses. In this study, 50 automated machine learning based probabilistic PT and study design taggers were built and applied to all articles in PubMed.

**Materials and Methods:** PubMed article metadata from 1987-2014 were used as training data, with 2015 used for recalibration. The set of articles indexed with a particular study design MeSH term or PT tag was used as positive training sets. For each PT, the rest of the literature from the same time period was used as its negative training set. Multiple features based on each article title, abstract and metadata were used in training the models. Taggers were evaluated on PubMed articles from 2016 and 2019. A manual analysis was also performed.

**Results:** Of the 50 predictive models that we created, 44 of these achieved an AUC of ∼0.90 or greater, with many having performance above 0.95. Of the clinically related study designs, the best performing was SYSTEMATIC_REVIEW with an AUC of 0.998; the lowest performing was RANDOM_ALLOCATION, with an AUC of 0.823.

**Discussion:** This work demonstrates that is feasible to build a large set of probabilistic publication type and study design taggers with high accuracy and ranking performance. Automated tagging permits users to identify qualifying articles as soon as they are published, and allows consistent criteria to be applied across different bibliographic databases. Probabilistic predictive scores are more flexible than binary yes/no predictions, since thresholds can be tailored for specific uses such as high recall literature search, user-adjustable retrieval size, and quality improvement of manually annotated databases.

**Conclusion:** The PT predictive probability scores for all PubMed articles are freely downloadable at http://arrowsmith.psych.uic.edu/evidence_based_medicine/mt_download.html for incorporation into user tools and workflows. Users can also perform PubMed queries at our Anne O’Tate value-added PubMed search engine http://arrowsmith.psych.uic.edu/cgi-bin/arrowsmith_uic/AnneOTate.cgi and filter retrieved articles according to both NLM-annotated and model-predicted publication types and study designs.

## BACKGROUND AND SIGNIFICANCE

The biomedical literature is often used to retrieve and synthesize knowledge for uses such as evidence-based medicine, systematic review, meta-analysis, practice guidelines, narrative reviews, and knowledge discovery. Identifying appropriate articles involves time consuming searching and manual review [1–8]. New computer-based tools and approaches are required to keep up with the ever-expanding biomedical literature base. Filtering literature based on study designs can provide substantial benefit early in the systematic review process [9]. One way to retrieve or filter literature is based on their indexing according to Medical Subject Headings (MeSH) [10]. Only a subset of MeSH headings refer to study designs or allocation schemes such as case-control, cross-over, and randomized clinical trials. Previous work on automated MeSH indexing of publications has primarily focused on predicting a small set of top or “best” set of MeSH terms for a given article. For example, the National Library of Medicine’s Medical Text Indexer (MTI) predicted a set of MeSH terms to be used as suggestions to a manual indexer [11]. Huang et al. [12] built a MeSH term recommender system that used PubMed Related Articles and a learning-to-rank approach. More recently, a combined MeSHLabeler [13] and DeepMeSH system used learning to rank along with Deep Learning semantic representation to achieve the highest binary prediction scores in the BioASQ2 and BioASQ3 challenges [14,15]. FullMeSH has enhanced these approaches by applying section-based convolutional neural networks to full article text [16]. MeSHProbeNet builds on these approaches, improving the time and space efficiency by creating fixed dimensional context vectors and predicting all MeSH terms simultaneously [17].

Previous work, by our team and others, has focused on using machine learning to rank or filter articles for inclusion in systematic reviews [18]. Tools such as RCT Tagger [19] and Robotsearch [20] allow identifying articles that are likely to be randomized controlled trials. Other tools such as RobotAnalyst [21], Abstracker [22], SWIFT-Review (https://www.sciome.com/swift-review/), and EPPI reviewer [23], incorporate machine learning processes to reduce the need for manual review based on initial groups of manual judgements.

Here our goal is also to create an automated MeSH indexing scheme for biomedical literature, but our approach differs in several important respects. First, in the present study, we focus only on those MeSH terms which refer to Publication Types (e.g., review, case report, news article, etc.) and clinically relevant study designs (e.g., case-control study, retrospective study, random allocation, etc.), both abbreviated here as PTs. Second, we do not attempt to index an article according to its “best” few MeSH terms, but rather consider each possible PT on its own merits, and attempt to estimate the probability that the article can be regarded as belonging to that PT. In previous work, we have proposed the use of automated assignment of probability confidence-based tags which provide a predictive score between 0.0 and 1.0 for randomized controlled trials and human-related studies created by machine learning models [19]. In the work presented here, a set of 50 probabilistic machine-learning based taggers have been developed and evaluated for a wide range of publication types and study designs, and applied to the entire PubMed indexed literature.

### OBJECTIVE

Build a large set of highly accurate probabilistic taggers for many of the most commonly used publication types and study designs. Provide a dataset of predictive scores for all PubMed articles that is publicly accessible and regularly incremented.

## MATERIALS AND METHODS

### Training data sets

To train, calibrate, and test the set of probabilistic tagger machine learning models, separate training, validation, and evaluation datasets were created, consisting of PubMed articles having abstracts and written in English (or, if not written in English, having English abstracts), which were human-related (i.e., were indexed with the *Humans* MeSH term). All qualifying PubMed articles published between 1987-2014 made up the *training* data set. This data set was used to select features, models, and algorithms. All qualifying PubMed articles published in 2015 made up the *validation* data set. This data set was used for initial testing and software verification, and also used to improve model calibration using a novel training and re-calibration approach that will be described later. Qualifying articles published in 2016 and 2019 made up the *evaluation* test data sets. These articles were kept separate until we had final models to test, therefore these articles were only used for final testing and performance evaluation. All of the final models were evaluated on the 2016 evaluation data set, except for SYSTEMATIC_REVIEW which was evaluated on 2019 article MEDLINE annotations (described later). The total training dataset contains almost 8 million publications (see Supplementary Table S1).

### Reference standards for systematic reviews and diagnostic test accuracy articles

By consensus within our group, which included two informaticians, a librarian, and a systematic review expert, we came up with a list of 55 of the most common and most needed publication types and study designs (PTs) used in EBM, systematic reviews, and other literature summarization and integration tasks, such as practice guidelines. For 54 of the 55 PTs that we initially examined, NLM has annotated formal Publication Type or MeSH terms for all articles indexed in MEDLINE. While it is well known that MEDLINE annotations are not perfect and that there is some error and inter-rater variability [24–27], overall the annotations are of very high quality. The annotations based on PT and MeSH terms were then used to both train the machine learning models as well as to perform automated evaluation of the models. Prior work has found that MEDLINE annotations provide enough resources to accurately train and evaluate probabilistic tagger models [19,20,28,29].

At the time this work was begun, there was no manually assigned MEDLINE annotation for SYSTEMATIC_REVIEW. This MeSH term was introduced starting with articles published in 2019. Articles published before 2019 have been assigned this MeSH term retroactively by NLM via an automated and non-probabilistic search process [30], and have not been manually reviewed by MEDLINE annotators (in contrast to new articles going forward). We found these retrospective annotations to be too noisy for our purposes in this work, and so we used a query-based method to generate a noisy training set of positive examples, and modified the machine learning approach slightly to take this into account. The PubMed query used to generate the positive training set for SYSTEMATIC_REVIEW was:

((((“systematic literature review” [tiab] OR “systematic review” [tiab] OR “search strategy” [tiab] OR “cochrane database syst rev” [ta]) AND (review[pt] OR meta-analysis[pt]) NOT (letter [pt] OR newspaper article [pt] OR comment [pt])))) Articles in the 2019 data set indexed by NLM with the SYSTEMATIC_REVIEW MeSH term were considered gold standard positive examples.

One other publication type we modeled also does not have a manual MeSH term assignment, DIAGNOSTIC_TEST_ACCURACY (DTA). This was also handled using a search process to build a noisy set of positive examples. The PubMed query used to generate the positive training set for DIAGNOSTIC_TEST_ACCURACY was:

~~~
((((“diagnostic accuracy” [ti] OR “diagnostic test accuracy” [ti]) NOT (letter
[pt] OR editorial [pt] OR review [pt] OR “practice guideline” [pt]))))
~~~

However, since there does not yet exist an acceptable gold standard corpus of DTA articles, or even an official consensus definition for DTAs, we will defer discussion and evaluation of this PT to a separate publication.

PubMed search strategies used to generate positive training sets for the other PTs are given in the Supplementary Appendix, Table S2.

### Representing each PT as a single vector

Our overall strategy of modeling proceeded as follows: Each of the PTs were represented as a reference cluster, i.e., as the set of articles in the training set indexed with that PT (possibly with some further restrictions as noted in Supplementary Table S2). Each article was represented as a multi-dimensional vector of feature scores (see below). Then, each PT was represented as a single multi-dimensional cluster vector of feature scores, by simply taking the average (or “centroid”) across all of the article vectors. To score a PubMed article in our scheme, we represent the article as a vector of feature scores, and calculate the similarity (i.e., inverse distance) between the article vector and each of the PT cluster vectors. The closer an article is to a given PT cluster, the more likely it is to belong to that PT [31]. The article citation-based feature scores are appended to the PT similarity scores and these combined feature vectors are fed into a support vector machine learning algorithm to compute the probability of an article for each of the 50 PTs.

### Feature set generation

Article titles and abstracts were converted to lower case text, normalized for ASCII characters and whitespace, stop words removed, and bigrams extracted as terms. Title bigrams and abstract bigrams were modeled separately as binary features. Our prior work on publication type tagging [19,28], and preliminary cross-validation work on the 2016 training data on a subset of these publication types found no improvement by including title or abstract term frequency information in these features. Stop words were removed using Andrew McCallum’s stop word list (available at http://mallet.cs.umass.edu/).

“Important word” cluster metric features were computed by counting the document frequency of all non-stop words in all of the article titles plus abstract in the cluster for that publication type. The top 100 most frequent words were then chosen to represent the publication type cluster. For an individual article the cluster similarity was computing by summing the weighted ranks for the top 100 cluster words that appear in the article title plus abstract. Words were weighted by the inverse rank using the formula w_i_ = 1 / (r_i_ + 1) where r_i_ is the rank in the top 100 word list for a word present in a specific article, and w_i_ is the weight added to the cluster similarity score. In this manner an important word similarity cluster score feature was computed for every article for every one of the 56 publication type clusters.

“Implicit word similarity” metric features were computed by used the implicit term vectors described in our prior published research [32]. Briefly, this approach looks at the terms that co-occur disproportionately with the terms actually contained within an article’s title plus abstract, using all of MEDLINE as the unsupervised corpus for computing term-term association within article titles and abstracts. The odds ratio of two terms appearing together in the unsupervised corpus is computed, and for each term the top 300 term by odds ratio are saved as the most related “implicit terms”. Then the publication type cluster “centroids” are computed in the following manner. For each unique term appearing in the combined titles and abstracts for all articles in a cluster, the log odds ratio for each term’s implicit terms are summed. Then the 300 implicit terms with the highest scoring log odds ratio sums are selected as the cluster implicit term “vector”. To compute the cluster similarity score of an individual article with a cluster implicit term vector, the article title and abstract top 300 implicit terms are computed in a similar manner as the cluster, and the fraction of overlapping implicit terms between the article and the cluster is used as the score.

“Embedded document vector similarity” cluster metric features were computed by first using Paragraph2Vec [33] to compute document vectors with 300 dimensions for every article in our data sets. The cluster vector centroids were computed for each publication type cluster by summing the document vectors for every article within a publication type cluster, and then normalizing this vector. Individual article cluster similarity scores were then computed simply by taking the dot product between the normalized cluster vector and the individual document vector. In this manner a Paragraph2Vec similarity score was computed for each document for each publication type cluster.

“Journal similarity” scores were computed by first analyzing the training data set for each journal and each publication type, computing the odds ratio of an article from a specific journal having a specific publication type as comparing to having that publication type regardless of where the article was published. To normalize these odds ratios to a useful range, the log base 100 of the odds ratio was computed and clipped to the interval [-1.0, +1.0]. For journals that did not include a minimum of 5 articles of a specific publication type, the normalized odds ratio of the most similar journal (as determined in separate work by our team based on MeSH profiling [34]) was used as long as the odds ratio was greater than one, otherwise a score of zero was assigned. For each publication type cluster, each article was assigned a similarity score based on the normalized odds of the article’s journal for the publication type.

In summary, the title and abstract bigram features included potentially millions of features each, and the cluster similarity feature sets (important words, implicit word similarity, embedded document vectors, and journal similarity) contained exactly 56 features each. All of these feature sets were used in the creation of all of the probabilistic taggers.

### Machine learning approach

The feature generation process resulted in a sparse article feature vector representing a large set of features in six classes: title bigrams, abstract bigrams, import words cluster metrics, implicit terms cluster metrics, Paragraph2Vec document cluster metrics, and journal cluster metrics. Preliminary testing applying cross-validation with the LibLinear implementation of SVM [35] on the 2016 training set and using five initial publication types (Case Control Studies, Cohort Studies, Cross-Over Studies, Cross-Sectional Studies, and Randomized Controlled Trials), showed that all feature classes were necessary to achieve maximum AUC when using both forward and backward feature class selection (as in our prior research [19]). Therefore, all feature classes were used in the creating the full models for the full set of publication types. The LibLinear classifier was chosen for this work because of its ability to handle large datasets and large numbers of features, and converge within our imposed time limit of 24 hours training per topic.

All of the publication type assignments present a highly unbalanced machine learning problem, as the tags are present in at most about 10% of articles (and usually much less). It is important to use all the positive sample data that this available so that it is not overshadowed by the overwhelming amount of negative sample data. For each of the PTs the classifier was trained on data consisting of all the positive examples in the 1987-2014 training set along with a random 20% sample of the negatives for that topic from the 1987-2014 training set. In other words, all of the in-cluster articles and a random selection of 20% of the out-of-cluster articles were used for training. This approach maximizes the use of the positive training samples and includes a representative sampling of negative examples up to what would fit into system random access memory in our 128GB computer that we used to process the data. One PT machine learning model was trained at a time, using the same set of n-gram and publication type similarity-based features as described above for every model. This approach also addresses another problem with highly unbalanced supervised learning, by ensuring that the prevalence of the positive samples is high enough to force the classification algorithm to make positive predictions at a useful rate, which might not happen if the algorithm can be 99% accurate by always predicting the negative class.

### Recalibration of estimated probabilities

Training the models in this way uses all of the positive examples, but presents a positive sample prevalence to the classifier that is much different from that actually found in the MEDLINE database. Therefore, predicted probabilities in the model will be incorrect, and the model calibration will highly skewed towards predicting high scores. To compensate for this, we included a post-processing recalibration step which was applied after the machine learning model was built. To recalibrate the models, isotonic regression was applied to the signed margin predictions from the liblinear SVM model, which was run on the 2015 data set to compute uncalibrated predictions and construct an isotonic regression model which transforms signed margin distance predictions into well-calibrated model probabilities between 0.0 and 1.0 that represent the frequency of the positive class in groups of samples having approximately the same output probability values. The sklearn.isotonic.IsotonicRegression module in scikit-learn (https://scikit-learn.org) was employed using a Python wrapper around liblinear to accomplish this.

As noted above, the machine learning models for SYSTEMATIC_REVIEW did not have human-generated training or recalibration data and were trained instead in the following manner. All of the articles in the cluster, which were identified by the publication type search were used as the positive training set, as well as a 20% sample of the negative articles. However, the n-grams present in the search terms (see above), were specifically elided from a random sample of the positive training data vectors. This was done to decrease the influence of the search terms in the machine learning process, and force the model to use give weight to other features. If the search terms were not removed from some of the training samples, then the machine learning model could perfectly predict the training data simply by highly weighting these few terms specified in the search and ignoring all the other features in the training vectors. On the other hand, if the search terms were removed from all of the positive training samples, then important n-gram features commonly present in unseen samples would be removed from the model, hurting performance. Preliminary testing on one year of the training data showed that the n-grams used in the search were down-weighted in the model when the search terms were elided from a randomly selected 20% of the SYSTEMATIC_REVIEW training samples. We used this level of term eliding when training the liblinear model for this publication type. Since recalibration data was not available, for this model, the Rüping-based method of normalizing the probabilistic predictions described in our prior work was used [19]. This provides a useful probabilistic prediction, albeit the Rüping method of normalization is typically not as good as a data-driven calibration approach such as isotonic regression.

In this manner, 50 publication type taggers (one for each of the types that had sufficient positive samples excluding DIAGNOSTIC_TEST_ACCURACY) were built using the machine learning approach described above. We did not create predictive scores for RETRACTION_OF_PUBLICATION, NEWPAPER_ARTICLE, SCIENTIFIC_INTEGRITY_REVIEW, and PUBLISHED_ERRATUM because there were too few positive samples. One of the PTs, CLINICAL_TRIAL, had a bug in the query during PT training set construction, and so we do not provide predictive scores for this PT. However, a predictive tag for CLINICAL_STUDY is included, which can be used instead.

CLINICAL_STUDY is a direct parent tag of CLINICAL_TRIAL in the MeSH hierarchy – that is, articles belonging to CLINICAL_STUDY should include both clinical trial articles as well as other Clinical Studies.

### Evaluation Approach

The publication type taggers were evaluated in several ways. For every tagger besides SYSTEMATIC_REVIEW, the 2016 data set, along with the corresponding gold standard MeSH terms and publication type tags, was used to do a large-scale automated evaluation. For SYSTEMATIC_REVIEW the manual MEDLINE annotations assigned in the year 2019 were used as the gold standard positive evaluation set with the negative set comprised of the rest of the literature from that year.

An extensive manual evaluation was also done for three of the most important publication types for systematic review: CROSS_SECTIONAL_STUDY, COHORT_STUDY, and CASE_CONTROL_STUDY. Similar manual evaluations have been done previously for RANDOMIZED_CONTROLLED_TRIAL in our prior work [19]. For this manual evaluation, cases of extreme disagreement between the probabilistic tagger prediction and the MEDLINE assigned MeSH tags were reviewed. These are the cases where the MeSH term HAS been assigned by MEDLINE yet the tagger score is very low, or the MeSH term has NOT been assigned, yet the tagger score is very high. These cases were collected by sorting the tagger predictions on the 2016 data set high to low, and then taking the first 100 articles sorted by predictive probability high to low that did not have the MeSH term assigned in MEDLINE, and then sorting low to high, and taking the first 100 articles with predictive probability scores < 0.10 that did have the MeSH term assigned in MEDLINE.

Manual judgments of PT assignments were made by independent reviewers who did not know, or consult, the MEDLINE indexing or the predicted tagger scores for the articles in question. Following a training phase on sample items, reviewers were given definitions based on and compatible with the MEDLINE definitions but enhanced with additional guidance for the reviewers (see final iterative updated definitions of CASE_CONTROL_STUDY, COHORT_STUDY, and CROSS_SECTIONAL_STUDY in Supplementary Table S3). Two reviewers independently read and marked 200 abstracts for each PT as meeting the definition for the publication type (“IS_PUBTYPE”), not meeting the definition of the publication type (“NOT_PUBTYPE”), or “UNCERTAIN”. When the abstract was unclear or did not give sufficient information, the reviewers had the option to view the full-text of the article, in which case they noted that they viewed the full-text and whether or not the full-text was essential for making a publication type determination. In a note field, reviewers indicated complexities of a given item. Then, a third person (JS) identified where the reviewers’ manual judgements agreed as well as disagreements and then met with the two reviewers to resolve disagreements during several reconciliation meetings to discuss each PT. In the reconciliation meetings, abstracts were re-read and discussed to determine the consensus determination of each item as meeting the definition for the publication type, not meeting the definition of the publication type, or “UNCERTAIN”. Conceptual differences, such as conflicting definitions, overlapping definitions, and closely related descriptions, can cause confusion in human or automated tagging and required the most discussion. An example is the difference between a cross-sectional <u>design</u> and a cross-sectional <u>study</u> (which uses a cross-sectional design but also requires selection from a population). Another source of ambiguity is that a single paper can use multiple study designs when it presents a series of experiments or analyses. Our approach is to assign a publication type if the corresponding design exists in the paper; and note that, with few exceptions (e.g., RCT and case control), the machine training sets had not excluded articles that described multiple methods. Complete details of the expert review process are given in Supplement S4.

## RESULTS

### Evaluation overview

Large-scale evaluation was performed comparing the predicted scores to gold standard MEDLINE assigned tags from future publication years collected after the training data. In addition, as described above, a detailed extreme disagreement manual review and analysis was done on the top 200 predictions that differ greatly from the MEDLINE assignments. The large-scale evaluation performance for each of the publication type taggers are shown in Tables 1 and 2. Table 1 contains the results for the clinically related PTs and study designs, and Table 2 contains results for the other PTs.

**Table 1.**
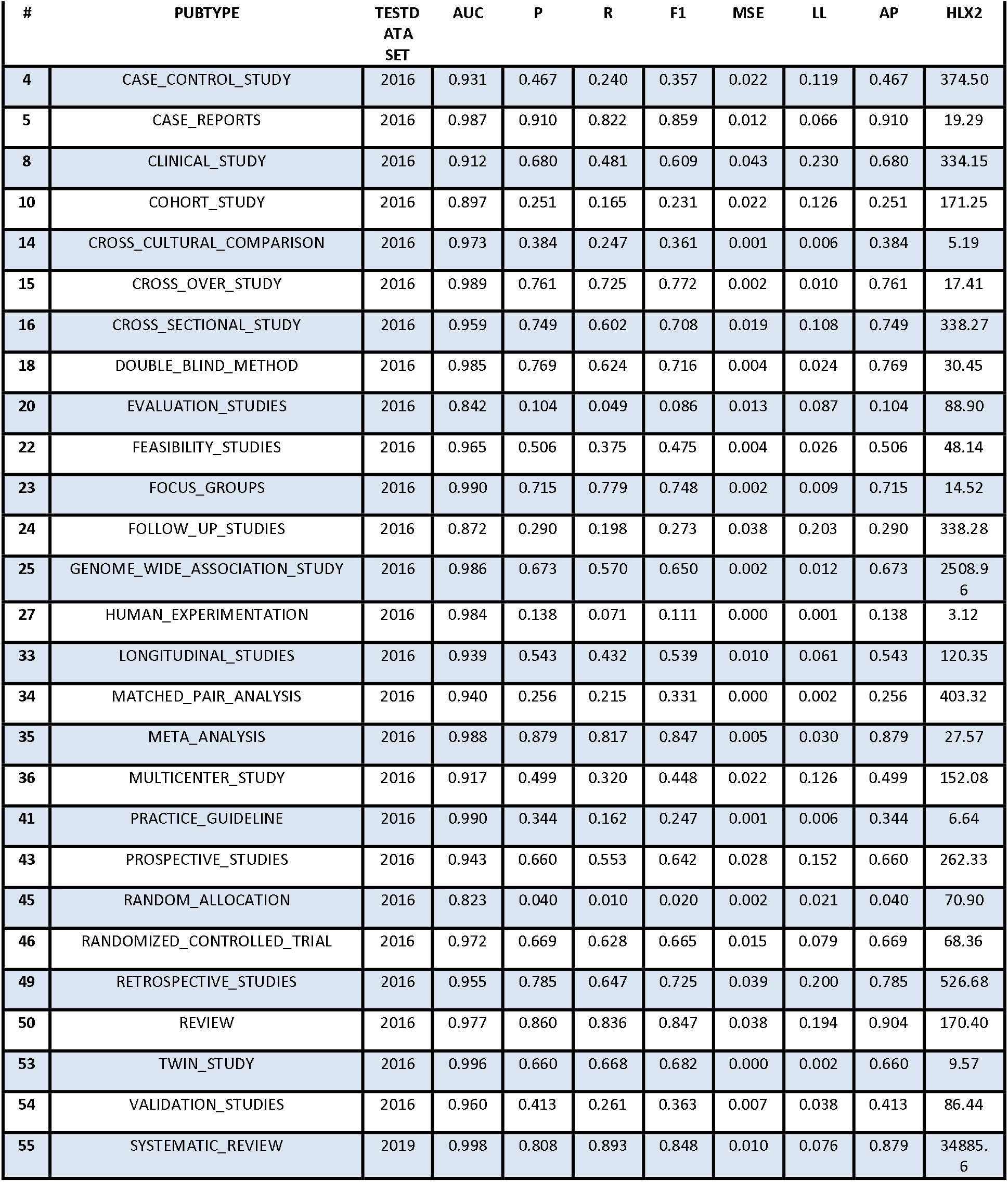
Tagger performance for the clinically related publication types and study designs based on the corresponding evaluation test sets detailed in Table 1. Several rank and probabilistic prediction accuracy-based measures are shown. These measures were all computed using the PubMed MeSH terms on articles having the *Humans* MeSH term as a gold standard. AUC = area under receiver operating curve, P = precision at 0.50 threshold, R = recall at 0.50 threshold, F1 = f-measure with beta = 1.0 at 0.50 threshold, MSE = mean squared error, LL = log loss, AP = average precision, HLX2 = Hosmer-Lemeshow Chi-Squared.

| # | PUBTYPE | TESTD<br>ATA<br>SET | AUC | P | R | F1 | MSE | LL | AP | HLX2 |
| --- | --- | --- | --- | --- | --- | --- | --- | --- | --- | --- |
| 4 | CASE_CONTROL_STUDY | 2016 | 0.931 | 0.467 | 0.240 | 0.357 | 0.022 | 0.119 | 0.467 | 374.50 |
| 5 | CASE_REPORTS | 2016 | 0.987 | 0.910 | 0.822 | 0.859 | 0.012 | 0.066 | 0.910 | 19.29 |
| 8 | CLINICAL_STUDY | 2016 | 0.912 | 0.680 | 0.481 | 0.609 | 0.043 | 0.230 | 0.680 | 334.15 |
| 10 | COHORT_STUDY | 2016 | 0.897 | 0.251 | 0.165 | 0.231 | 0.022 | 0.126 | 0.251 | 171.25 |
| 14 | CROSS_CULTURAL_COMPARISON | 2016 | 0.973 | 0.384 | 0.247 | 0.361 | 0.001 | 0.006 | 0.384 | 5.19 |
| 15 | CROSS_OVER_STUDY | 2016 | 0.989 | 0.761 | 0.725 | 0.772 | 0.002 | 0.010 | 0.761 | 17.41 |
| 16 | CROSS_SECTIONAL_STUDY | 2016 | 0.959 | 0.749 | 0.602 | 0.708 | 0.019 | 0.108 | 0.749 | 338.27 |
| 18 | DOUBLE_BLIND_METHOD | 2016 | 0.985 | 0.769 | 0.624 | 0.716 | 0.004 | 0.024 | 0.769 | 30.45 |
| 20 | EVALUATION_STUDIES | 2016 | 0.842 | 0.104 | 0.049 | 0.086 | 0.013 | 0.087 | 0.104 | 88.90 |
| 22 | FEASIBILITY_STUDIES | 2016 | 0.965 | 0.506 | 0.375 | 0.475 | 0.004 | 0.026 | 0.506 | 48.14 |
| 23 | FOCUS_GROUPS | 2016 | 0.990 | 0.715 | 0.779 | 0.748 | 0.002 | 0.009 | 0.715 | 14.52 |
| 24 | FOLLOW_UP_STUDIES | 2016 | 0.872 | 0.290 | 0.198 | 0.273 | 0.038 | 0.203 | 0.290 | 338.28 |
| 25 | GENOME_WIDE_ASSOCIATION_STUDY | 2016 | 0.986 | 0.673 | 0.570 | 0.650 | 0.002 | 0.012 | 0.673 | 2508.96 |
| 27 | HUMAN_EXPERIMENTATION | 2016 | 0.984 | 0.138 | 0.071 | 0.111 | 0.000 | 0.001 | 0.138 | 3.12 |
| 33 | LONGITUDINAL_STUDIES | 2016 | 0.939 | 0.543 | 0.432 | 0.539 | 0.010 | 0.061 | 0.543 | 120.35 |
| 34 | MATCHED_PAIR_ANALYSIS | 2016 | 0.940 | 0.256 | 0.215 | 0.331 | 0.000 | 0.002 | 0.256 | 403.32 |
| 35 | META_ANALYSIS | 2016 | 0.988 | 0.879 | 0.817 | 0.847 | 0.005 | 0.030 | 0.879 | 27.57 |
| 36 | MULTICENTER_STUDY | 2016 | 0.917 | 0.499 | 0.320 | 0.448 | 0.022 | 0.126 | 0.499 | 152.08 |
| 41 | PRACTICE_GUIDELINE | 2016 | 0.990 | 0.344 | 0.162 | 0.247 | 0.001 | 0.006 | 0.344 | 6.64 |
| 43 | PROSPECTIVE_STUDIES | 2016 | 0.943 | 0.660 | 0.553 | 0.642 | 0.028 | 0.152 | 0.660 | 262.33 |
| 45 | RANDOM_ALLOCATION | 2016 | 0.823 | 0.040 | 0.010 | 0.020 | 0.002 | 0.021 | 0.040 | 70.90 |
| 46 | RANDOMIZED_CONTROLLED_TRIAL | 2016 | 0.972 | 0.669 | 0.628 | 0.665 | 0.015 | 0.079 | 0.669 | 68.36 |
| 49 | RETROSPECTIVE_STUDIES | 2016 | 0.955 | 0.785 | 0.647 | 0.725 | 0.039 | 0.200 | 0.785 | 526.68 |
| 50 | REVIEW | 2016 | 0.977 | 0.860 | 0.836 | 0.847 | 0.038 | 0.194 | 0.904 | 170.40 |
| 53 | TWIN_STUDY | 2016 | 0.996 | 0.660 | 0.668 | 0.682 | 0.000 | 0.002 | 0.660 | 9.57 |
| 54 | VALIDATION_STUDIES | 2016 | 0.960 | 0.413 | 0.261 | 0.363 | 0.007 | 0.038 | 0.413 | 86.44 |
| 55 | SYSTEMATIC_REVIEW | 2019 | 0.998 | 0.808 | 0.893 | 0.848 | 0.010 | 0.076 | 0.879 | 34885.6 |

**Table 2.**
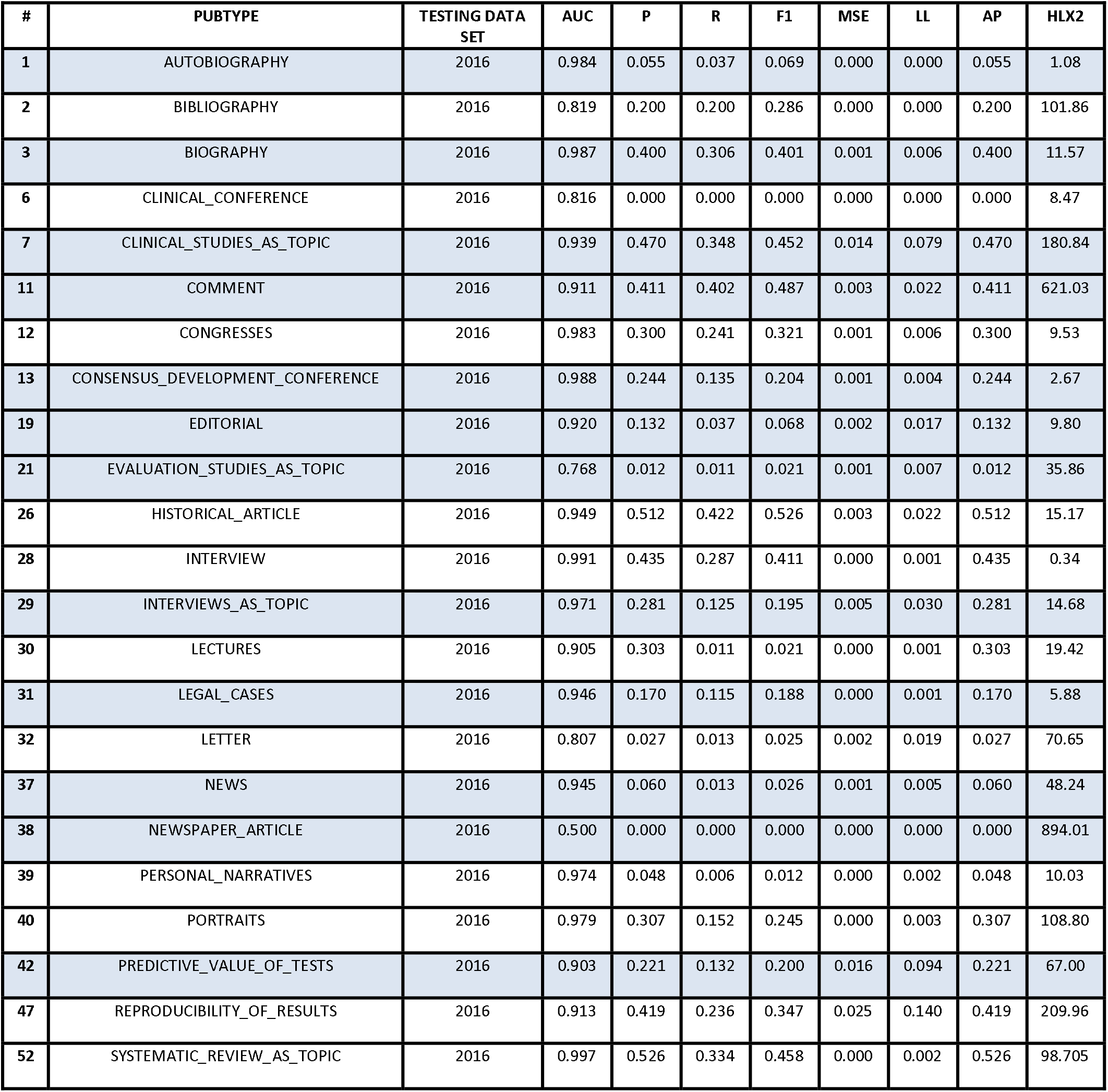
Tagger performance for the other publication types and study designs used to create the cluster-based features. Several rank and probabilistic prediction accuracy-based measures are shown. based on the corresponding evaluation test sets detailed in Table 1. These measures were all computed using the PubMed MeSH terms on articles having the *Humans* MeSH term as a gold standard. AUC = area under receiver operating curve, P = precision at 0.50 threshold, R = recall at 0.50 threshold, F1 = f-measure with beta = 1.0 at 0.50 threshold, MSE = mean squared error, LL = log loss, AP = average precision, HLX2 = Hosmer-Lemeshow Chi-Squared.

| # | PUBTYPE | TESTING DATA SET | AUC | P | R | F1 | MSE | LL | AP | HLX2 |
| --- | --- | --- | --- | --- | --- | --- | --- | --- | --- | --- |
| 1 | AUTOBIOGRAPHY | 2016 | 0.984 | 0.055 | 0.037 | 0.069 | 0.000 | 0.000 | 0.055 | 1.08 |
| 2 | BIBLIOGRAPHY | 2016 | 0.819 | 0.200 | 0.200 | 0.286 | 0.000 | 0.000 | 0.200 | 101.86 |
| 3 | BIOGRAPHY | 2016 | 0.987 | 0.400 | 0.306 | 0.401 | 0.001 | 0.006 | 0.400 | 11.57 |
| 6 | CLINICAL_CONFERENCE | 2016 | 0.816 | 0.000 | 0.000 | 0.000 | 0.000 | 0.000 | 0.000 | 8.47 |
| 7 | CLINICAL_STUDIES_AS_TOPIC | 2016 | 0.939 | 0.470 | 0.348 | 0.452 | 0.014 | 0.079 | 0.470 | 180.84 |
| 11 | COMMENT | 2016 | 0.911 | 0.411 | 0.402 | 0.487 | 0.003 | 0.022 | 0.411 | 621.03 |
| 12 | CONGRESSES | 2016 | 0.983 | 0.300 | 0.241 | 0.321 | 0.001 | 0.006 | 0.300 | 9.53 |
| 13 | CONSENSUS_DEVELOPMENT_CONFERENCE | 2016 | 0.988 | 0.244 | 0.135 | 0.204 | 0.001 | 0.004 | 0.244 | 2.67 |
| 19 | EDITORIAL | 2016 | 0.920 | 0.132 | 0.037 | 0.068 | 0.002 | 0.017 | 0.132 | 9.80 |
| 21 | EVALUATION_STUDIES_AS_TOPIC | 2016 | 0.768 | 0.012 | 0.011 | 0.021 | 0.001 | 0.007 | 0.012 | 35.86 |
| 26 | HISTORICAL_ARTICLE | 2016 | 0.949 | 0.512 | 0.422 | 0.526 | 0.003 | 0.022 | 0.512 | 15.17 |
| 28 | INTERVIEW | 2016 | 0.991 | 0.435 | 0.287 | 0.411 | 0.000 | 0.001 | 0.435 | 0.34 |
| 29 | INTERVIEWS_AS_TOPIC | 2016 | 0.971 | 0.281 | 0.125 | 0.195 | 0.005 | 0.030 | 0.281 | 14.68 |
| 30 | LECTURES | 2016 | 0.905 | 0.303 | 0.011 | 0.021 | 0.000 | 0.001 | 0.303 | 19.42 |
| 31 | LEGAL_CASES | 2016 | 0.946 | 0.170 | 0.115 | 0.188 | 0.000 | 0.001 | 0.170 | 5.88 |
| 32 | LETTER | 2016 | 0.807 | 0.027 | 0.013 | 0.025 | 0.002 | 0.019 | 0.027 | 70.65 |
| 37 | NEWS | 2016 | 0.945 | 0.060 | 0.013 | 0.026 | 0.001 | 0.005 | 0.060 | 48.24 |
| 38 | NEWSPAPER_ARTICLE | 2016 | 0.500 | 0.000 | 0.000 | 0.000 | 0.000 | 0.000 | 0.000 | 894.01 |
| 39 | PERSONAL_NARRATIVES | 2016 | 0.974 | 0.048 | 0.006 | 0.012 | 0.000 | 0.002 | 0.048 | 10.03 |
| 40 | PORTRAITS | 2016 | 0.979 | 0.307 | 0.152 | 0.245 | 0.000 | 0.003 | 0.307 | 108.80 |
| 42 | PREDICTIVE_VALUE_OF_TESTS | 2016 | 0.903 | 0.221 | 0.132 | 0.200 | 0.016 | 0.094 | 0.221 | 67.00 |
| 47 | REPRODUCIBILITY_OF_RESULTS | 2016 | 0.913 | 0.419 | 0.236 | 0.347 | 0.025 | 0.140 | 0.419 | 209.96 |
| 52 | SYSTEMATIC_REVIEW_AS_TOPIC | 2016 | 0.997 | 0.526 | 0.334 | 0.458 | 0.000 | 0.002 | 0.526 | 98.705 |

### Large-scale automated evaluation

The AUC value provides an objective measure of how well a model can distinguish positive from negative examples. A perfect AUC is 1.0, whereas an AUC value of 0.5 indicates no discriminative value at all. As shown in Table 1, the clinically related publication types, which are commonly used in evidence-based medicine, had consistently high AUC values, for example: CASE_CONTROL_STUDY (0.931), CASE_REPORTS (0.987), COHORT_STUDY (0.897), CROSS_OVER_STUDY (0.989), CROSS_SECTIONAL_STUDY (0.959), META_ANALYSIS (0.988), RANDOMIZED_CONTROLLED_TRIAL (0.972), and RETROSPECTIVE_STUDIES (0.955). Although the model was trained exclusively on human-related articles, we found that the predictive performance was generally similar when tested on articles lacking *Humans* MeSH indexing (shown in Supplement Table S5). Calibration of the predictive scores was also evaluated. The calibration of the tagger prediction values is also very good, as demonstrated by the overall low values of the Hosmer-Lemeshow metric. This means that after recalibration, the model predictive scores have a clear and accurate probabilistic interpretation [19].

Table 2 shows the evaluation results for the other PTs. Performance on AUC ranges from a high of 0.988 for CONSENSUS_DEVELOPMENT_CONFERENCE, to a low of 0.768 for EVALUATION_STUDIES_AS_TOPIC. The wide variety of performance scores in this set is due to several factors. First, some of the PTs are very rare, having relatively few samples in both the training and evaluation data sets, relative to the PTs shown in Table 1. Furthermore, some of these PTs, such as CLINICAL_CONFERENCE and LETTER are very heterogeneous and can contains a wide variety of scientific topics and methods. Combining few samples with heterogeneous topics results in very sparse training data. The calculated precision, recall, and F1 scores are based on converting predictive scores to binary yes/no assignments based on 0.5 as a default threshold value.

### RCT Tagger vs. Multi-tagger scores for Randomized Controlled Trials

Although Multi-tagger employed different feature sets and ML methods for tagging randomized controlled trials than our previous RCT Tagger tool [19], the performance of both models was quite similar. Using the set of 2018 PubMed articles as a test set, we found that RCT Tagger tended to give scores that were only slightly higher than the corresponding Multi-tagger score (mean difference = 0.023, 95% CI of the mean difference = (0.0229, 0.0233)). The AUC of Multi-tagger = 0.963, which is almost identical to that of RCT Tagger = 0.966 tested on the same articles. We did observe that an ensemble prediction, taking the maximum score across both models, produced a still better AUC = 0.972, and so the ensemble model may be preferred in cases where the best possible recall is required.

### Independent manual evaluation

Our team experts conducted an independent formal review and consensus process for extreme disagreements between MEDLINE indexing and model predictive scores for CASE_CONTROL_STUDY, COHORT_STUDY, and CROSS_SECTIONAL_STUDY publication types (see Supplementary Table S4 and the Dryad data repository file). When our tagger gave a high predictive score (>0.9) on articles that lacked a corresponding MEDLINE indexing term, independent review suggested that the model assignment was correct in almost all cases (CROSS_SECTIONAL_STUDY (99%), CASE_CONTROL_STUDY (94.9%), and COHORT STUDY (92.2%)). Conversely, when articles received MEDLINE indexing but model predictive scores were very low (<0.1), independent review suggested that the model assignment was correct in the majority of cases: CASE_CONTROL_STUDY (85.4%), COHORT STUDY (76.3%), and CROSS_SECTIONAL_STUDY (53.6%). Note that according to the MeSH database, CROSS_SECTIONAL_STUDY refers to “Studies in which the presence or absence of disease or other health-related variables are determined *<u>in each member of the study population or in a representative sample</u>* at one particular time.” (our emphasis). The relatively low agreement between MEDLINE indexing practice and model scores for the PT CROSS_SECTIONAL_STUDY may, at least in part, reflect the fact that many PubMed articles incorporate a cross-sectional design (i.e., comparing variables at one particular time), which may lead some indexers to mark such articles even if they fail to satisfy the more stringent sampling requirements to qualify them as true cross-sectional studies.

Also notable is the extremely high performance of the SYSTEMATIC_REVIEW tagger (AUC = 0.99; Table 1). Binary classification measures for this tagger on the 2019 evaluation data were Precision=0.808, Recall=0.893, F1 = 0.848. This is especially encouraging given that the training data was based on search criteria only and the test data consisted of more recent 2019 literature manually annotated by NLM.

### Implementation

Using the models developed above, a vector of 50 predictive scores, one for each PT, was created for every article in PubMed and stored in a database. This database is regularly updated; we plan on daily updates. Two interfaces for accessing these results have also been created: First, the predictive scores can be freely downloaded as bulk data files from http://arrowsmith.psych.uic.edu/evidence_based_medicine/mt_download.html and integrated into user specified workflows and applications. Second, we have derived a secondary dataset in which the predictive scores (estimated probabilities between 0 and 1) have been converted into binary yes/no predictions for each PT, by choosing threshold values that are optimized for F1 for each PT, or in other words, the threshold values that exhibit the best balance between recall and precision for each PT. Using these binary PT assignments, we have integrated the secondary dataset into our Anne O’Tate search tool [36], which allows slicing and dicing the search retrieval in many different ways, including publication types. Users can find additional predicted PubMed articles for a PT, which may include new articles not yet MEDLINE-indexed, as well as articles indexed by PubMed that are not contained in MEDLINE, and those that might have been missed by MEDLINE indexing processes (Figure 1). Note that the predicted articles may outnumber the MEDLINE-indexed articles for a given query (Figure 1). Precision, recall, and F1 performance metrics for the binary thresholds used by Anne O’Tate are shown in Table 3.

**Table 3.**
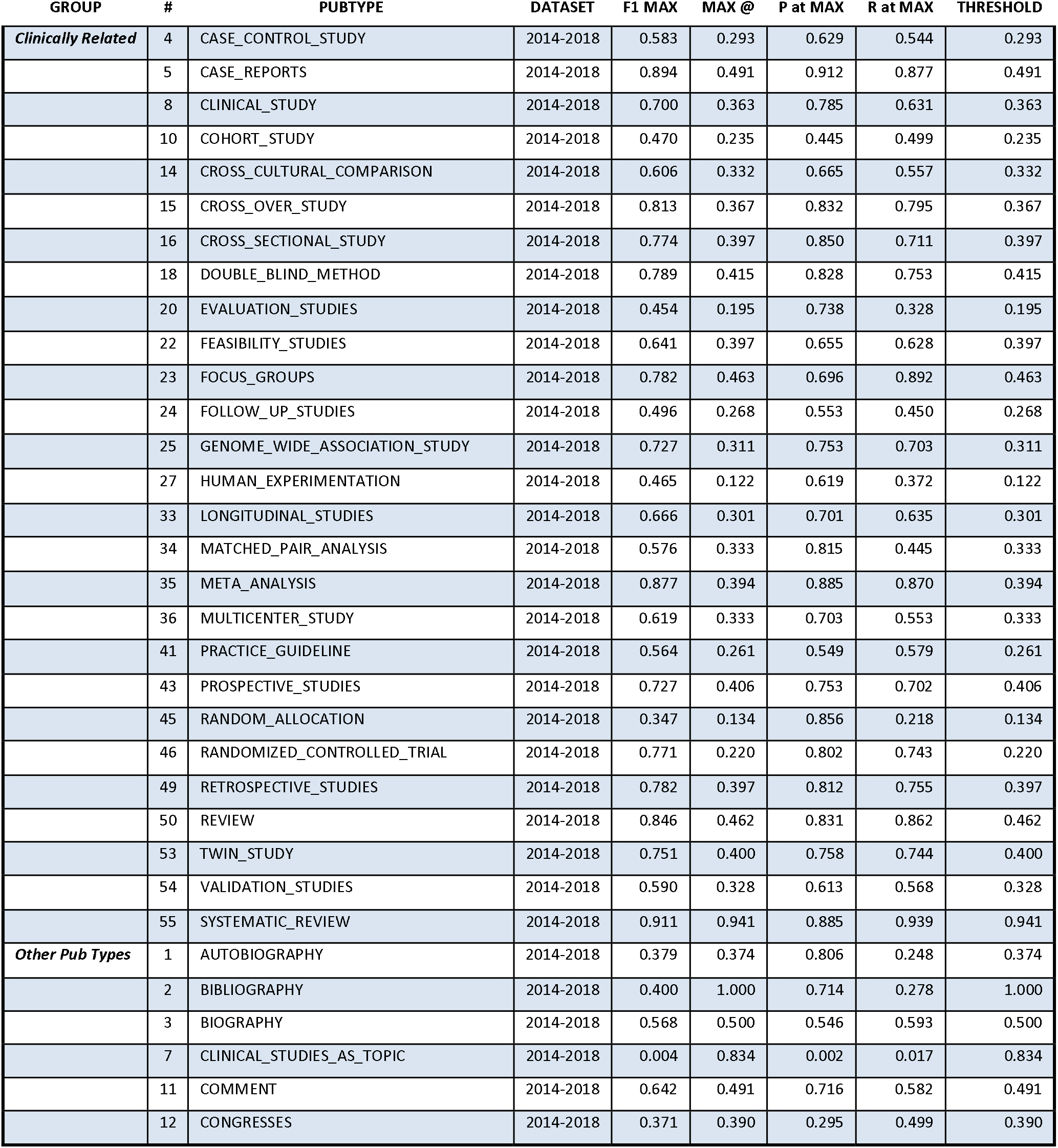
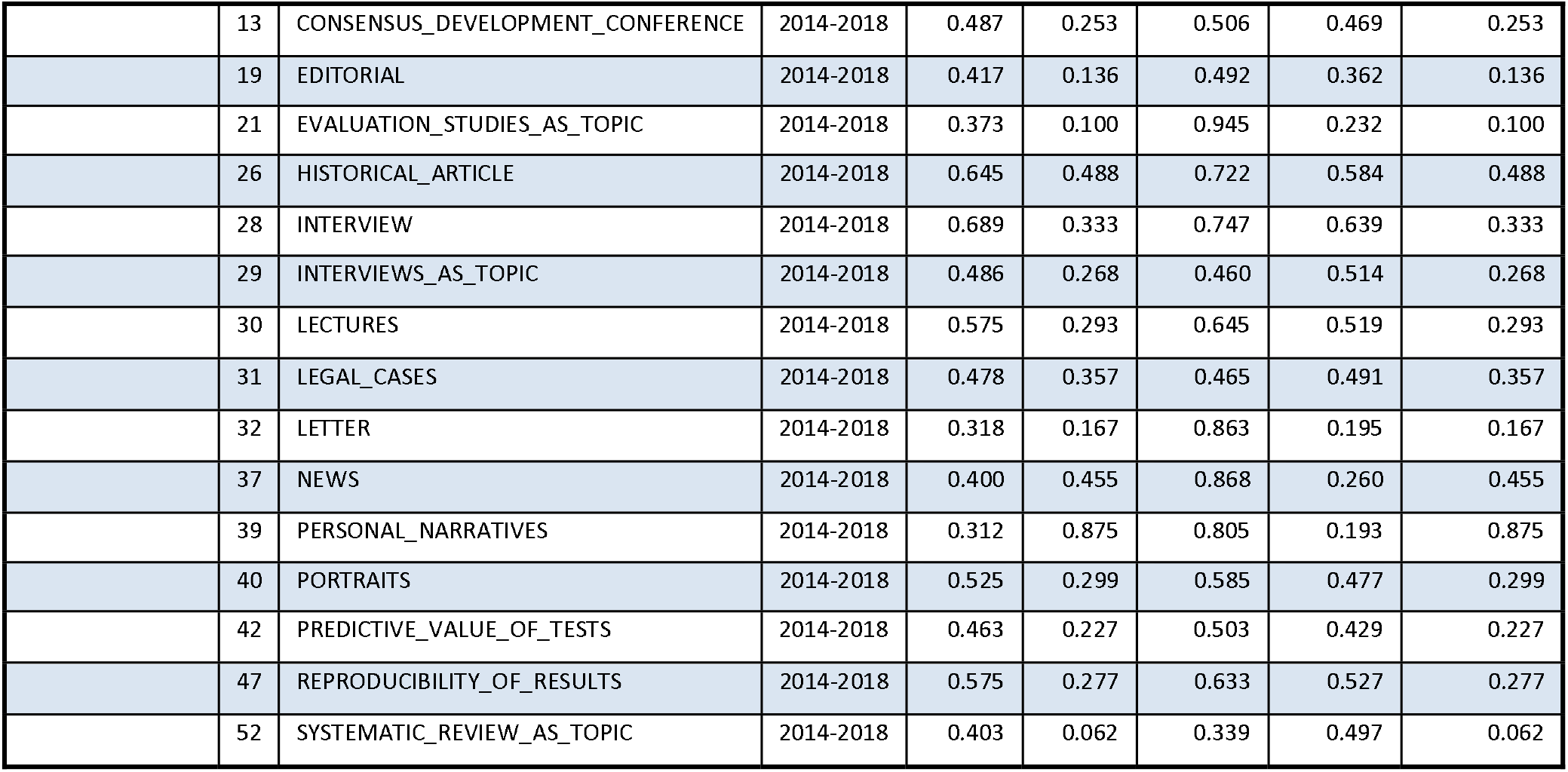
Tagger model thresholds for designation of predicted positive cases used in Anne O’Tate interface. These thresholds were computed using the PubMed MeSH terms on articles having the *Humans* MeSH term as a gold standard for publication years 2014 to 2018, inclusive. The threshold for Diagnostic Test Accuracy was derived from a sample of 1,000 EMBASE articles. F1 MAX = maximum F1 accuracy achieved across all possible thresholds from [0,1]. MAX @ = model score where F1 MAX occurs. P at MAX = precision at F1 MAX threshold, R at MAX = recall at F1 MAX threshold. THRESHOLD = model score threshold for designating a positive case.

| GROUP | # | PUBTYPE | DATASET | F1 MAX | MAX @ | P at MAX | R at MAX | THRESHOLD |
| --- | --- | --- | --- | --- | --- | --- | --- | --- |
| *Clinically Related* | 4 | CASE_CONTROL_STUDY | 2014-2018 | 0.583 | 0.293 | 0.629 | 0.544 | 0.293 |
|  | 5 | CASE_REPORTS | 2014-2018 | 0.894 | 0.491 | 0.912 | 0.877 | 0.491 |
|  | 8 | CLINICAL_STUDY | 2014-2018 | 0.700 | 0.363 | 0.785 | 0.631 | 0.363 |
|  | 10 | COHORT_STUDY | 2014-2018 | 0.470 | 0.235 | 0.445 | 0.499 | 0.235 |
|  | 14 | CROSS_CULTURAL_COMPARISON | 2014-2018 | 0.606 | 0.332 | 0.665 | 0.557 | 0.332 |
|  | 15 | CROSS_OVER_STUDY | 2014-2018 | 0.813 | 0.367 | 0.832 | 0.795 | 0.367 |
|  | 16 | CROSS_SECTIONAL_STUDY | 2014-2018 | 0.774 | 0.397 | 0.850 | 0.711 | 0.397 |
|  | 18 | DOUBLE_BLIND_METHOD | 2014-2018 | 0.789 | 0.415 | 0.828 | 0.753 | 0.415 |
|  | 20 | EVALUATION_STUDIES | 2014-2018 | 0.454 | 0.195 | 0.738 | 0.328 | 0.195 |
|  | 22 | FEASIBILITY_STUDIES | 2014-2018 | 0.641 | 0.397 | 0.655 | 0.628 | 0.397 |
|  | 23 | FOCUS_GROUPS | 2014-2018 | 0.782 | 0.463 | 0.696 | 0.892 | 0.463 |
|  | 24 | FOLLOW_UP_STUDIES | 2014-2018 | 0.496 | 0.268 | 0.553 | 0.450 | 0.268 |
|  | 25 | GENOME_WIDE_ASSOCIATION_STUDY | 2014-2018 | 0.727 | 0.311 | 0.753 | 0.703 | 0.311 |
|  | 27 | HUMAN_EXPERIMENTATION | 2014-2018 | 0.465 | 0.122 | 0.619 | 0.372 | 0.122 |
|  | 33 | LONGITUDINAL_STUDIES | 2014-2018 | 0.666 | 0.301 | 0.701 | 0.635 | 0.301 |
|  | 34 | MATCHED_PAIR_ANALYSIS | 2014-2018 | 0.576 | 0.333 | 0.815 | 0.445 | 0.333 |
|  | 35 | META_ANALYSIS | 2014-2018 | 0.877 | 0.394 | 0.885 | 0.870 | 0.394 |
|  | 36 | MULTICENTER_STUDY | 2014-2018 | 0.619 | 0.333 | 0.703 | 0.553 | 0.333 |
|  | 41 | PRACTICE_GUIDELINE | 2014-2018 | 0.564 | 0.261 | 0.549 | 0.579 | 0.261 |
|  | 43 | PROSPECTIVE_STUDIES | 2014-2018 | 0.727 | 0.406 | 0.753 | 0.702 | 0.406 |
|  | 45 | RANDOM_ALLOCATION | 2014-2018 | 0.347 | 0.134 | 0.856 | 0.218 | 0.134 |
|  | 46 | RANDOMIZED_CONTROLLED_TRIAL | 2014-2018 | 0.771 | 0.220 | 0.802 | 0.743 | 0.220 |
|  | 49 | RETROSPECTIVE_STUDIES | 2014-2018 | 0.782 | 0.397 | 0.812 | 0.755 | 0.397 |
|  | 50 | REVIEW | 2014-2018 | 0.846 | 0.462 | 0.831 | 0.862 | 0.462 |
|  | 53 | TWIN_STUDY | 2014-2018 | 0.751 | 0.400 | 0.758 | 0.744 | 0.400 |
|  | 54 | VALIDATION_STUDIES | 2014-2018 | 0.590 | 0.328 | 0.613 | 0.568 | 0.328 |
|  | 55 | SYSTEMATIC_REVIEW | 2014-2018 | 0.911 | 0.941 | 0.885 | 0.939 | 0.941 |
| *Other Pub Types* | 1 | AUTOBIOGRAPHY | 2014-2018 | 0.379 | 0.374 | 0.806 | 0.248 | 0.374 |
|  | 2 | BIBLIOGRAPHY | 2014-2018 | 0.400 | 1.000 | 0.714 | 0.278 | 1.000 |
|  | 3 | BIOGRAPHY | 2014-2018 | 0.568 | 0.500 | 0.546 | 0.593 | 0.500 |
|  | 7 | CLINICAL_STUDIES_AS_TOPIC | 2014-2018 | 0.004 | 0.834 | 0.002 | 0.017 | 0.834 |
|  | 11 | COMMENT | 2014-2018 | 0.642 | 0.491 | 0.716 | 0.582 | 0.491 |
|  | 12 | CONGRESSES | 2014-2018 | 0.371 | 0.390 | 0.295 | 0.499 | 0.390 |
|  | 13 | CONSENSUS_DEVELOPMENT_CONFERENCE | 2014-2018 | 0.487 | 0.253 | 0.506 | 0.469 | 0.253 |
|  | 19 | EDITORIAL | 2014-2018 | 0.417 | 0.136 | 0.492 | 0.362 | 0.136 |
|  | 21 | EVALUATION_STUDIES_AS_TOPIC | 2014-2018 | 0.373 | 0.100 | 0.945 | 0.232 | 0.100 |
|  | 26 | HISTORICAL_ARTICLE | 2014-2018 | 0.645 | 0.488 | 0.722 | 0.584 | 0.488 |
|  | 28 | INTERVIEW | 2014-2018 | 0.689 | 0.333 | 0.747 | 0.639 | 0.333 |
|  | 29 | INTERVIEWS_AS_TOPIC | 2014-2018 | 0.486 | 0.268 | 0.460 | 0.514 | 0.268 |
|  | 30 | LECTURES | 2014-2018 | 0.575 | 0.293 | 0.645 | 0.519 | 0.293 |
|  | 31 | LEGAL_CASES | 2014-2018 | 0.478 | 0.357 | 0.465 | 0.491 | 0.357 |
|  | 32 | LETTER | 2014-2018 | 0.318 | 0.167 | 0.863 | 0.195 | 0.167 |
|  | 37 | NEWS | 2014-2018 | 0.400 | 0.455 | 0.868 | 0.260 | 0.455 |
|  | 39 | PERSONAL_NARRATIVES | 2014-2018 | 0.312 | 0.875 | 0.805 | 0.193 | 0.875 |
|  | 40 | PORTRAITS | 2014-2018 | 0.525 | 0.299 | 0.585 | 0.477 | 0.299 |
|  | 42 | PREDICTIVE_VALUE_OF_TESTS | 2014-2018 | 0.463 | 0.227 | 0.503 | 0.429 | 0.227 |
|  | 47 | REPRODUCIBILITY_OF_RESULTS | 2014-2018 | 0.575 | 0.277 | 0.633 | 0.527 | 0.277 |
|  | 52 | SYSTEMATIC_REVIEW_AS_TOPIC | 2014-2018 | 0.403 | 0.062 | 0.339 | 0.497 | 0.062 |

**Figure 1.**
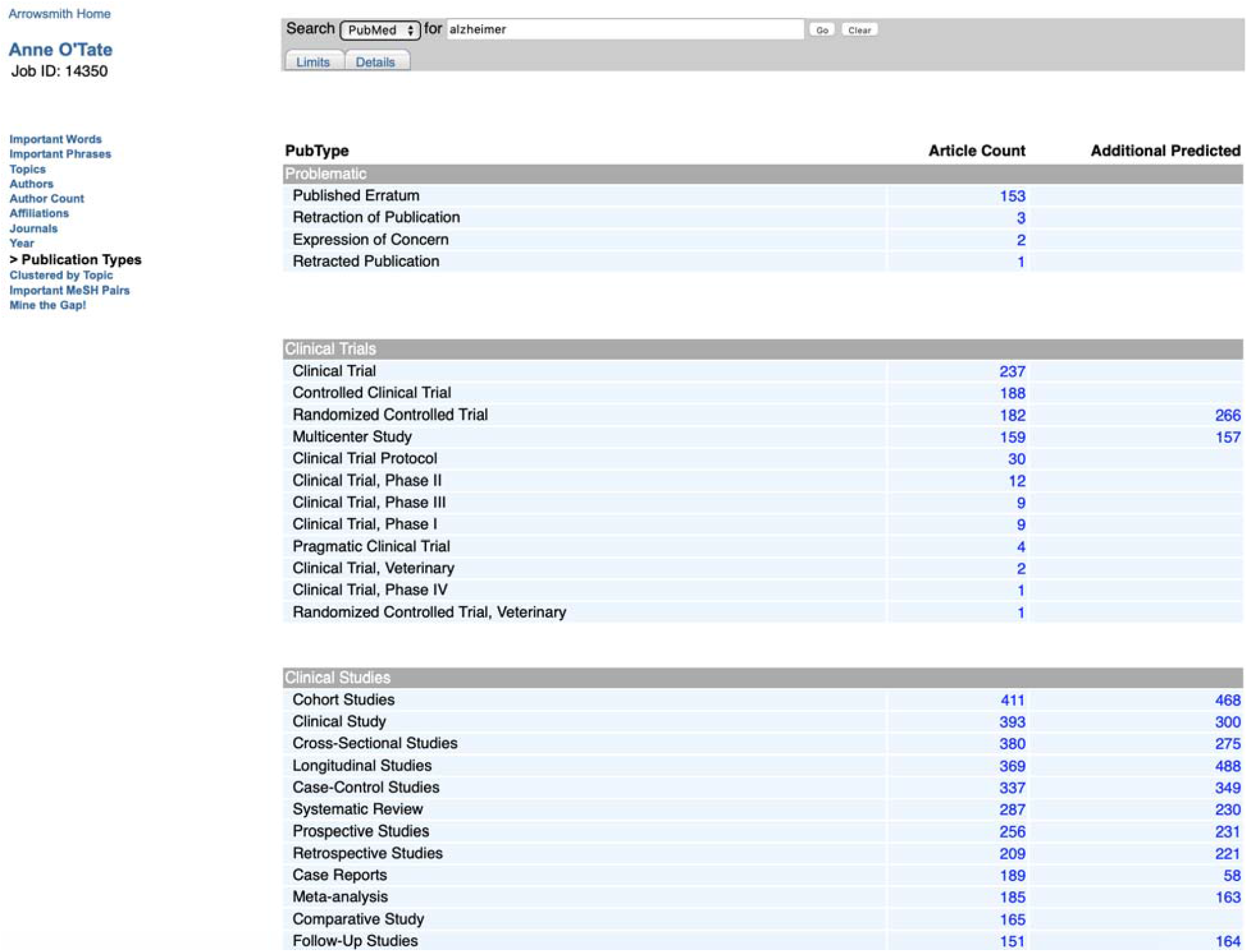
A query on “Alzheimer” in PubMed retrieves a large set of articles that are displayed by PT in the Anne O’Tate “Publication Types” tool. This page shows “Article Count” which displays the MEDLINE-indexed articles for each PT, as well as “Additional Predicted” articles predicted by Multi-tagger.

## DISCUSSION

We have built a large set of 50 probabilistic publication type and study design taggers with good accuracy and ranking performance. These predictive scores can be generated automatically and quickly, as soon as an article is published. This can be a powerful tool to aid in information retrieval and filtering of the biomedical literature and increase the timeliness of these tasks.

Probabilistic publication type taggers provide several advantages over prior approaches:

1. Articles can be filtered with an adjustable threshold. This allows tailoring the size of the returned filtered set to be appropriate to the size of the topic literature base and available review resources. Furthermore, different use cases can apply customized thresholds to better reflect their needs. For example, when high recall is essential, as in retrieving relevant RCT articles for systematic reviews, we have previously shown that discarding articles having very low RCT predictive scores (<0.01) can remove the majority of non-RCT articles while maintaining extremely high recall (99.4%) [37].
2. Scores can be applied consistently to articles indexed in multiple databases. Because the automated approach proposed here only requires citation information common to all literature databases, the tags can be applied to an article no matter what database(s) in which it appears. In principle, searches can be conducted and filtered across databases and ranked and combined using a uniform process.
3. MEDLINE and other bibliographic databases could potentially use the predictive scores as a prompt to assist manual indexing, or to identify possible indexing errors.
4. Not all articles are either 100% or 0% of a PT. It may be that an article partially satisfies PT criteria enough to be of interest to some users—for example, a paper with a cross-sectional design, but that does not meet the sampling requirements to be a cross-sectional study.
5. Rapid review groups could consider using low Multi-tagger scores to “filter out” items, when MeSH-based PTs are too noisy or when they face extreme resource constraints. In our research, currently we are testing various filtering strategies using the taggers against published systematic reviews, starting from the key questions, PTs of interest for each key question, and their actual search results. Our goal is to estimate the work savings that the tools can provide at a given level of recall.
6. Other potentially powerful applications include combining taggers to rank on multiple criteria, and pooling tagger predictions to estimate the size of a literature evidence base. Taggers could be combined by multiplying probabilities to perform an “AND” operation for narrowing selection of literature, for example for use cases requiring studies to be both RCTs and cross-over studies.

A potential limitation of the Multi-tagger model is that most, if not all, PTs represent a small fraction of the entire literature, and at the same time, positive gold standard examples exhibit a wide range of predictive scores; this means that for any given threshold, a significant number of “negative” examples will be predicted as having that PT. We have minimized false positives by recalibrating predictive scores to take imbalanced data into account, and users can further minimize false positives by setting high thresholds. However, our examination of extreme disagreements with MEDLINE indexing indicates that articles receiving high predictive scores generally do satisfy the criteria for that PT. Hence, our model may help identify relevant articles even within the “negative” population, as well as among articles which did not receive MEDLINE indexing because they are newly published or because they are in journals not indexed by MEDLINE.

## CONCLUSION

Multi-tagger represents a new approach to automatic indexing of the biomedical literature. Besides facilitating the retrieval and filtering of articles according to publication type and study design by end-users, the predictive scores should be valuable as features to be incorporated into larger automated machine learning-based efforts. For example, our work is complementary to other efforts to automatically assign MeSH terms to articles [10-16]; one might extend the Multi-tagger approach to predicting other types of MeSH terms, or alternatively, predicted PT scores may potentially improve the accuracy of other machine learning approaches [10-16]. Another potential use of Multi-tagger is to assist in automated ranking of articles for inclusion in systematic reviews [6,24,25]. Simply screening for publication type has substantial value for identifying relevant articles for inclusion, [24, 37]. Yet publication type tagging is complementary to the use of textual or semantic similarity measures to predict relevance for inclusion (e.g., risk of bias [39]), so one would expect that publication type tagging will help improve the performance of other existing machine learning approaches. Therefore, the Multi-tagger dataset should represent a valuable resource not only for end-users but for the biomedical text mining community.

## Data Availability

The PT predictive probability scores for all PubMed articles are freely downloadable at http://arrowsmith.psych.uic.edu/evidence_based_medicine/mt_download.html for incorporation into user tools and workflows. Users can also perform PubMed queries at our Anne O'Tate value-added PubMed search engine http://arrowsmith.psych.uic.edu/cgi-bin/arrowsmith_uic/AnneOTate.cgi and filter retrieved articles according to both NLM-annotated and model-predicted publication types and study designs.
Furthermore, the data underlying the Manual Review of Extreme Disagreements will be deposited and made available in the Dryad Digital Repository.

## Acknowledgements

The authors wish to thank Luesoni Johnson Kuck and Yu ‘Amber’ Wang for annotating articles and participating in expert review consensus.

## Funding Statement

This work was supported by National Institutes of Health (NIH)/National Library of Medicine grant number R01LM010817.

## Data Availability Statement

The data underlying the Manual Review of Extreme Disagreements will be available in the Dryad Digital Repository

Multi-Tagger data produced in this paper are available for download at http://arrowsmith.psych.uic.edu/evidence_based_medicine/mt_download.html

## Competing Interests Statement

The authors have no competing interests to declare.

## Contributorship Statement

The conceptualization, methodology, and supervision of this project was primarily performed by AMC and NRS with domain expert input from MM. The software was primarily written by AMC with contributions from AWH. AMC, JS, YF, PD, AWH, and NRS performed the data curation, formal analysis and validation of the results. The original draft was primarily written by AMC, JS, and NRS. All authors contributed to the review and final editing of the manuscript for publication.

## SUPPLEMENTAL APPENDIX

**Supplementary Table S1.**
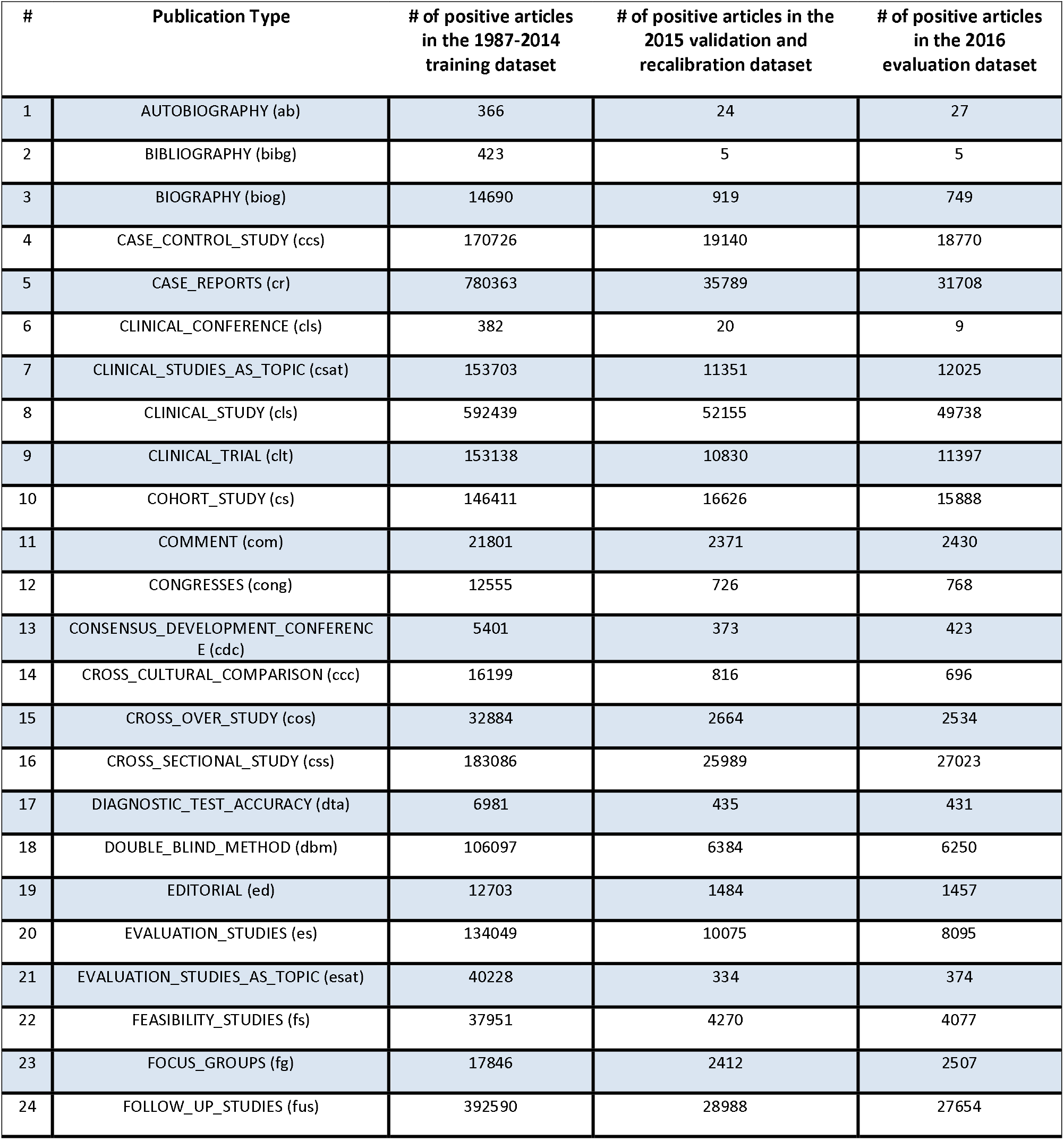
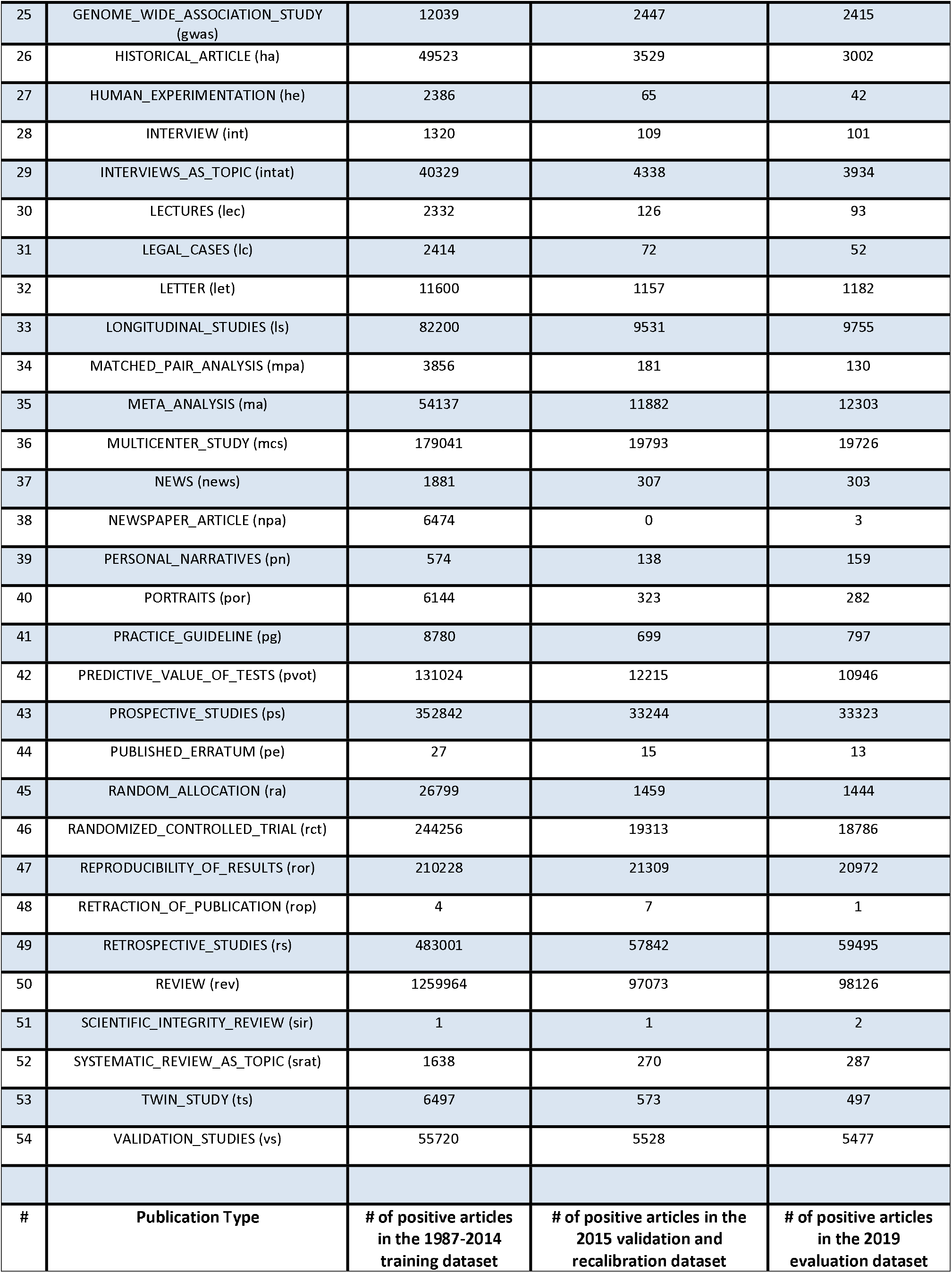
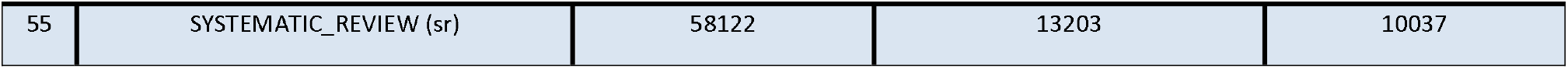
Composition of each of the data sets used in this work, including total number of positive articles in each data set for each publication type. For SYSTEMATIC_REVIEW the evaluation data set is based on articles published in 2019, for all publication types, the evaluation data set is based on articles published in 2016. There were a total of 7,976,632 articles in the 1987-2014 training set, 601,422 articles in the 2015 validation/recalibration data set, 594,101 articles in the 2016 evaluation data set, and 288,345 articles in the 2019 evaluation data set.

**Supplementary Table S2**. List of queries used to define each PT in the training sets. This is not simply having a given Mesh term or PT, but may include some restrictions as well.

### Publication Types and Search Strategies

All search strategies assume limiting years, MeSH Humans tag, [hasabstract] (English Language OR English Abstract), adding to search strategy in this form:

“humans”[MeSH Terms]

AND (“1987”[PDAT] : “2014”[PDAT])

AND hasabstract[All Fields]

AND (“english”[Language] OR “english abstract”[Publication Type])

#### Publication Type Positive Training Set Searches

- Randomized Controlled Trials
  - Include: randomized controlled trial [pt]
  - Exclude: editorial[pt], letter[pt], practice guideline[pt], “case-control studies”[MeSH Terms], “cohort studies”[MeSH Terms], review[pt]
- Cross-Over Study
  - Include: cross-over study[mh]
  - Exclude: editorial[pt], letter[pt], practice guideline[pt], review[pt]
- Cohort Study
  - Include: cohort studies[mh]
  - Exclude: editorial[pt], letter[pt], practice guideline[pt], “case-control studies”[MeSH Terms], review[pt], clinical trial [pt]
- Case-Control Study
  - Include: “case-control studies”[MeSH Terms]
  - Exclude: editorial[pt], letter[pt], practice guideline[pt], “cohort studies”[MeSH Terms], review[pt], clinical trial [pt]
- Cross-Sectional Study
  - Include: cross-sectional study [mh]
  - Exclude: editorial[pt], letter[pt], practice guideline[pt], review[pt]
- Diagnostic Test Accuracy Study
  - (SEARCH: `((((“diagnostic accuracy” [ti] OR “diagnostic test accuracy” [ti]) NOT (letter [pt] OR editorial [pt] OR review [pt] OR “practice guideline” [pt]))))`
- Meta-Analysis
  - (SEARCH: Meta-Analysis [Publication Type] NOT Meta-Analysis as Topic[mh])
- Case Reports
  - (SEARCH: “Case Reports” [Publication Type] NOT Editorials [Publication Type])
- Clinical Conference
  - (SEARCH Clinical Conference [Publication Type] NOT Case Reports [Publication Type])
- Letter
  - (SEARCH: Letter [Publication Type])
- Editorial
  - (SEARCH: Editorial [Publication Type])
- Comment
  - (SEARCH: Comment [Publication Type])
- Multicenter Study
  - (SEARCH: Multicenter Study [Publication Type])
- Practice Guideline
  - (SEARCH: “practice guideline”[Publication Type] NOT “practice guidelines as topic”[MeSH Terms]
- Review
  - (SEARCH: Review [Publication Type])
- Autobiography
  - (SEARCH: Autobiography [Publication Type])
- Published Erratum
  - (SEARCH “Published Erratum”[Publication Type])
- Retraction of Publication
  - (SEARCH: “Retraction of Publication”[Publication Type])
- Prospective studies
  - (SEARCH: “Prospective studies”[mh:noexp])
- Retrospective studies
  - (SEARCH: “Retrospective studies”[mh:noexp])
- Longitudinal Studies
  - (SEARCH: “Longitudinal Studies”[mh:noexp])
- Follow-Up Studies
  - (SEARCH: “Follow-Up Studies”[mh:noexp])
- Genome-Wide Association Study
  - (SEARCH: “genome-wide association study”[mh])
- Focus Groups
  - (SEARCH: “Focus Groups”[mh:noexp])
- Feasibility Studies
  - (SEARCH: “Feasibility Studies”[mh:noexp])
- Cross-Cultural Comparison
  - (SEARCH: “Cross-Cultural Comparison”[mh])
- Reproducibility of Results
  - (SEARCH: “Reproducibility of Results”[mh])
- Double-Blind Method
  - (SEARCH: “Double-Blind Method”[mh:noexp])
- Matched-Pair Analysis
  - (SEARCH: “Matched-Pair Analysis”[mh:noexp])
- Random Allocation
  - (SEARCH: “Random Allocation”[mh:noexp])
- Human Experimentation
  - (SEARCH: “Human Experimentation”[mh:noexp])
- Interviews as Topic
  - (SEARCH: “Interviews as Topic” [mh:noexp])
- Systematic Review as Topic
  - (SEARCH: “systematic review or systematic reviews”[all fields] AND “Review Literature as Topic”[mh])
- Twin Study
  - (SEARCH: “Twin Study” [Publication Type])
- Bibliography
  - (SEARCH: Bibliography [Publication Type])
- Biography
  - (SEARCH: Biography [Publication Type])
- Congresses
  - (SEARCH: Congresses [Publication Type])
- Historical article
  - (SEARCH: “Historical article” [Publication Type])
- Portraits
  - (SEARCH: Portraits [Publication Type])
- Consensus Development Conference
  - (SEARCH: “Consensus Development Conference”[Publication Type])
- Lectures
  - (SEARCH: Lectures [Publication Type])
- Legal Cases
  - (SEARCH: “Legal Cases” [Publication Type])
- Personal narratives
  - (SEARCH “Personal narratives” [Publication Type])
- News
  - (SEARCH: News [Publication Type])
- Newspaper Article
  - (SEARCH: “Newspaper Article” [Publication Type])
- Interview [Publication Type]
  - (SEARCH: Interview [Publication Type])
- Clinical Study
  - (SEARCH: “Clinical Study” [Publication Type])
- Clinical Studies as Topic
  - (SEARCH: “Clinical Studies as Topic” [mh])
- Clinical Trial
  - (SEARCH: “Clinical Trial” [mh:exp])
- Evaluation Studies
  - (SEARCH: “Evaluation Studies” [Publication Type])
- Evaluation Studies as Topic
  - (SEARCH: “Evaluation Studies as Topic”[mh:noexp])
- Validation Studies
  - (SEARCH: “Validation Studies” [Publication Type])
- Scientific Integrity Review
  - (SEARCH: “Scientific Integrity Review” [Publication Type])
- Expression of Concern [Publication Type]
  - (SEARCH: “Expression of Concern” [Publication Type])
- Published Erratum
  - (SEARCH: “Published Erratum” [Publication Type])
- Retraction of Publication
  - (SEARCH: “Retraction of Publication” [Publication Type])
- Predictive Value of Tests[mh:noexp]
  - (SEARCH: “Predictive Value of Tests”[mh:noexp])
- Systematic Review
  - (SEARCH: ((((“systematic literature review” [tiab] OR “systematic review” [tiab] OR “search strategy” [tiab] OR “cochrane database syst rev” [ta]) AND (review[pt] OR meta-analysis[pt]) NOT (letter [pt] OR newspaper article [pt] OR comment [pt])))))

**Supplementary Table S3**

Expert Review Pubtype Operational Definitions with Application Notes

### Case-control study (from https://community.cochrane.org/glossary#letter-C)

A study that compares people with a specific disease or outcome of interest (cases) to people from the same population without that disease or outcome (controls), and which seeks to find associations between the outcome and prior exposure to particular risk factors. This design is particularly useful where the outcome is rare and past exposure can be reliably measured. Case-control studies are usually retrospective, but not always.

CLARIFICATION 1: A case control study can collect subjects (cases and controls) either retrospectively or prospectively, but cannot follow an outcome prospectively.

CLARIFICATION 2: A case control study may compare cases vs. controls by e.g., measuring current levels of molecules in the blood, carrying out MRI scans, obtaining genotypes, or examining test results. It is not necessary that things are examined that occurred in the past (prior to the subjects being identified as cases and controls). Thus, a case control study may also be a cross-sectional study (where subjects are collected and measured at the same time).

NOTE: Don’t trust a study’s assertion that it is a case control study: check what it does.

NOTE: Case control studies must be observational studies: a case control study cannot measure the outcome of an intervention.

NOTE: There must be a specified method or criteria for selecting cases and controls. NOTE: Usually there is some way of matching for confounds, but not always.

NOTE: Case control studies may compare genetic, environmental, lifestyle, and/or medical histories of cases and controls.

### Cohort study (from https://community.cochrane.org/glossary#letter-C)

“An **observational study** in which a defined group of people (the cohort) is followed over time. The **outcomes** of people in subsets of this cohort are compared, to examine people who were exposed or not exposed (or exposed at different levels) to a particular **intervention** or other factor of interest. A **prospective** cohort study assembles **participants** and follows them into the future. A **retrospective** (or historical) cohort study identifies subjects from past records and follows them from the time of those records to the present. Because subjects are not allocated by the investigator to different interventions or other exposures, **adjusted analysis** is usually required to minimise the influence of other factors (**confounders**).”

NOTE: A cohort study can just follow a population over time to see what distinguishes those who do vs. don’t get a disease or outcome.

NOTE: Many cohort studies are designed to examine a question about burden of disease (e.g. prevalence, incidence, mean levels).

NOTE: We consider “interventions” to include OTHER FACTORS OF INTEREST, for instance exposure to known things.

NOTE: The cohort that is studied can be made up of individual people, or other units such as schools, hospitals, prisons, or other groups of people.

NOTE: Don’t worry too much about how/whether dropouts are adjusted for. The last sentence of the definition gives advice for <u>quality</u> and interpretation of results: “Because subjects are not allocated by the investigator to different interventions or other exposures, adjusted analysis is usually required to minimise the influence of other factors (confounders).”

### Cross-sectional study (from https://community.cochrane.org/glossary#letter-C)

“A study measuring the distribution of some characteristic(s) in a population at a particular point in time.”

NOTE: Cross-sectional studies are descriptive studies (neither longitudinal nor experimental). They involve data collected at (approximately) the same time.

NOTE: “A population” could be a representative randomly sampled subset of the population (it is not necessary to have measurements on the entire population).

NOTE: Cross-sectional studies often measure prevalence or absolute risk of a condition in a population, or measure associations between risk factors and outcomes viewed at the same point in time.

NOTE: In a cross-sectional study, time cannot be a variable of interest: Whatever is studied has to be conceivable as a “moment in time”. So, a comparison between cross-sectional studies is NOT a cross-sectional study.

NOTE: For an article to “be” a study design, only one of those components has to meet the definition. So, a series of cross-sectional studies is considered cross-sectional because of any of the individual studies in the series.

**Supplement S4**

### Manual Review of Extreme Disagreements

Our extreme disagreement analysis uncovered several patterns and groups of errors for each of the three publication types reviewed, by rereading the abstracts for each False Positive (FP) and False Negative (FN) (see Dryad data repository file).

For case control errors, the most prominent pattern was that 14 FNs did not have typical case and/or control terminology in the abstract. For many of these, cases and controls were clear to humans based on domain knowledge, for instance “582 patients with chronic respiratory diseases and chronic non-respiratory diseases” (PMID:27358182). Other FNs used atypical phrasings: “The sample included 145 adolescents aged 13-17 years, 40% with exposure to child abuse.” (PMID:25979082). Full text was required in some cases. For example, the fact that samples were taken from cases and controls was only clear in full-text for PMID 26704906. The second most frequent error came from 6 FNs that were tissue studies, and their wording was different from that of clinical studies, e.g.: “We obtained 228 HCC samples from patients who underwent liver resection, 168 paired non-tumor adjacent cirrhotic liver samples, and 10 non-tumor liver tissues from patients undergoing resection for hepatic hemangioma.” (PMID: 27614046). Future systems could integrate more elaborate natural language processing techniques to identify the contrast between people with and without a condition. Tissue studies use significantly different language, which a tissue-specific subtype of case control studies might ameliorate.

In the cohort studies the most prominent pattern (8 FNs) used previous data from databases, chart reviews, and large-scale demographic surveys. Their descriptions of the data sources demarcate them from other cohort studies. For example, “In this study, we identified clinical risk factors for recurrent AKI present during index AKI hospitalizations that occurred between 2003 and 2010 using a regional Veterans Administration database in the United States.” (PMID: 26264853); “Data used in this study came from different waves of Demographic and Health Surveys (DHS) of Sub-Saharan countries.” (PMID: 27442118). The second most prominent category affected all 7 FPs, as the word “cohort” appeared in all 7 FP abstracts and the studies were related to cohort studies in some way. Examples include cross-sectional studies (PMID: 27150640, 26847995), an RCT (PMID: 27084245) done on a cohort and using baseline data from a cohort for analysis (PMID: 28185591), and a database for cohort studies (PMID: 27439404). Two categories affected 6 FNs each. In the first category, follow-up is unstated in the abstract or must be inferred by humans. For example, in (PMID: 27389407), follow-up is only stated in the full-text. Follow-up can also be stated in a less conventional way, such as “All patients were evaluated at baseline (prior to radiotherapy) and at 1 and 4 months after radiotherapy and then yearly thereafter for pain assessment using a visual analogue scale (VAS) and to determine the administration of analgesic treatment.” (PMID: 27885851). The second category uses cohort data to build risk models. In such abstracts, the content organization is different in that it emphasises the risk model rather than the study (e.g., PMID: 26683498). Cohort studies based on previous data or for building risk models can be treated in the future as finer-grained publication types, and we expect these complex FP cases to require human experts to screen them out. Unconventional or missing follow-up information will be tricky to deal with. One potential solution could be to build models that do not rely on methods information, such as training models by using abstract segments that report results.

For the cross-sectional errors, the most prominent pattern was 13 FNs that are genetics or biomarker studies, which measure genetics or biomarker characteristics in a population. They cause errors in the tagger because of the unusual wording. Example FNs: “We initially compared the predictive value of MK with other urinary biomarkers, including N-acetyl-β-D-glucosaminidase (NAG), interleukin (IL)-18, and neutrophil gelatinase-associated lipocalin (NGAL), for the detection and differential diagnosis of established AKI (549 patients).” (PMID: 27530994); “A total of 109 suspected isolates were characterized, but only 57 V. cholerae strains could be confirmed using multiplex real-time PCR as well as rpoB sequencing and typed as V. cholerae O:1 Ogawa biotype El Tor “ (PMID: 27487957). Another prominent category (11 FNs) is cross-sectional case-control studies, in which subjects were chosen as cases or controls. They are not canonical cross-sectional studies, whose hallmark is random sampling in a population. However, our experts applied a less stringent definition and counted case-control studies that used a cross-sectional design (e.g., measurements were taken at one time) as cross-sectional studies. Likewise, the PubMed database also indexes some studies with both “Case-Control Studies” and “Cross-Sectional Studies” MeSH indexing terms. For another category (11 FNs), time was involved in some way. In such cases, the cross-sectional data were collected over a period rather than at once. For example, in order to study incidence of pertussis in an African population, “Mother-infant pairs were enrolled at 1 week of life, and then seen at 2-to 3-week intervals through 14 weeks of age.” (PMID:27838668). Lastly, 9 FNs report clinical assessment or diagnostic tests, which come with cross-sectional data measured by such tests (e.g., PMID: 27374234). In the future, those categories could also be easier to detect as their own finer-grained types.

One policy question is whether or not to include systematic reviews that contained an instance of the PT being annotated as an example of that type. Upon a review of the error analysis, we found that we did not use a consistent policy between publication types. We annotated systematic reviews that contained case control studies as case control studies but did not annotate systematic reviews that included cross-sectional studies as cross-sectional studies. Systematic reviews account for 2 errors deemed FPs in the case control studies (PMID: 27604213, 25218914) and one error deemed a FN in the cross-sectional studies (PMID: 26987556). Since evidence synthesis groups may use systematic reviews to identify additional potentially relevant studies, reviews that include a particular study design may be relevant. In future work, the systematic review tagger could seek to classify whether or not particular study designs are included in a given systematic review.

**Supplementary Table S5.** Comparison of Multi-tagger performance on articles tagged with the MeSH term *Humans* vs. those not tagged as human articles. All articles having any assigned publication types were included from the years 2014-2018 in this evaluation. F1 at the optimal threshold is shown, along with precision and recall at that threshold. Area under the receiver operating curve (AUC) is also shown.

| Pubtype |  | Human |  |  |  |  |  |  | Non-Human |  |  |  |  |  |  |
| --- | --- | --- | --- | --- | --- | --- | --- | --- | --- | --- | --- | --- | --- | --- | --- |
| # | model | articles scored | # Positive by MeSH | F1 max | threshold at max | precision at F1max | recall at F1max | AUC | articles scored | # Positive by MeSH | F1 max | max occurs at | precision at F1max | recall at F1max | AUC |
| 1 | AUTOBIOGRAPHY | 2,534,711 | 117 | 0.379 | 0.374 | 0.806 | 0.248 | 0.981 | 1,251,934 | 98 | 0.219 | 0.082 | 0.176 | 0.289 | 0.972 |
| 2 | BIBLIOGRAPHY | 2,534,711 | 18 | 0.400 | 1.000 | 0.714 | 0.278 | 0.873 | 1,251,934 | 6 | 0.000 | 0.000 | 0.000 | 0.200 | 0.793 |
| 3 | BIOGRAPHY | 2,534,711 | 2,163 | 0.568 | 0.500 | 0.546 | 0.593 | 0.997 | 1,251,934 | 1,407 | 0.660 | 0.240 | 0.616 | 0.711 | 0.997 |
| 4 | CASE CONTROL STUDY | 2,534,711 | 83,795 | 0.583 | 0.293 | 0.629 | 0.544 | 0.934 | 1,251,934 | 4,324 | 0.151 | 0.318 | 0.113 | 0.230 | 0.875 |
| 5 | CASE REPORTS | 2,534,711 | 144,208 | 0.894 | 0.491 | 0.912 | 0.877 | 0.991 | 1,251,934 | 8,775 | 0.436 | 0.282 | 0.319 | 0.690 | 0.981 |
| 7 | CLINICAL STUDIES AS TOPIC | 2,534,711 | 288 | 0.004 | 0.834 | 0.002 | 0.017 | 0.880 | 1,251,934 | 18 | 0.002 | 0.655 | 0.001 | 0.500 | 0.992 |
| 8 | CLINICAL STUDY | 2,534,711 | 205,842 | 0.700 | 0.363 | 0.785 | 0.631 | 0.928 | 1,251,934 | 116 | 0.004 | 0.791 | 0.002 | 0.232 | 0.867 |
| 10 | COHORT STUDY | 2,534,155 | 79,790 | 0.470 | 0.235 | 0.445 | 0.499 | 0.906 | 1,251,540 | 3863 | 0.072 | 0.225 | 0.042 | 0.269 | 0.863 |
| 11 | COMMENT | 2,534,711 | 11,151 | 0.642 | 0.491 | 0.716 | 0.582 | 0.958 | 1,251,934 | 5,079 | 0.615 | 0.296 | 0.684 | 0.559 | 0.969 |
| 12 | CONGRESSES | 2,534,711 | 1,452 | 0.371 | 0.390 | 0.295 | 0.499 | 0.983 | 1,251,934 | 235 | 0.397 | 0.361 | 0.331 | 0.495 | 0.986 |
| 13 | CONSENSUS DEVELOPMENT CONFERENCE | 2,534,711 | 1,781 | 0.487 | 0.253 | 0.506 | 0.469 | 0.989 | 1,251,934 | 67 | 0.127 | 0.160 | 0.073 | 0.500 | 0.979 |
| 14 | CROSS CULTURAL COMPARISON | 2,534,711 | 3,058 | 0.606 | 0.332 | 0.665 | 0.557 | 0.980 | 1,251,934 | 172 | 0.023 | 0.083 | 0.012 | 0.286 | 0.945 |
| 15 | CROSS OVER STUDY | 2,534,711 | 11,040 | 0.813 | 0.367 | 0.832 | 0.795 | 0.987 | 1,251,934 | 1299 | 0.498 | 0.367 | 0.400 | 0.659 | 0.963 |
| 16 | CROSS SECTIONAL STUDY | 2,534,711 | 117,248 | 0.774 | 0.397 | 0.850 | 0.711 | 0.964 | 1,251,934 | 6,536 | 0.265 | 0.397 | 0.177 | 0.527 | 0.955 |
| 18 | DOUBLE BLIND METHOD | 2,534,711 | 26,919 | 0.789 | 0.415 | 0.828 | 0.753 | 0.987 | 1,251,934 | 1284 | 0.178 | 0.415 | 0.113 | 0.418 | 0.929 |
| 19 | EDITORIAL | 2,534,711 | 6,293 | 0.417 | 0.136 | 0.492 | 0.362 | 0.927 | 1,251,934 | 778 | 0.229 | 0.126 | 0.231 | 0.227 | 0.857 |
| 20 | EVALUATION STUDIES | 2,534,711 | 36,009 | 0.454 | 0.195 | 0.738 | 0.328 | 0.880 | 1,251,934 | 0 | 0.142 | 0.078 | 0.113 | 0.192 | 0.812 |
| 21 | EVALUATION STUDIES AS TOPIC | 2,534,711 | 1,556 | 0.373 | 0.100 | 0.945 | 0.232 | 0.797 | 1,251,934 | 239 | 0.034 | 0.039 | 0.056 | 0.024 | 0.755 |
| 22 | FEASIBILITY STUDIES | 2,534,711 | 17,602 | 0.641 | 0.397 | 0.655 | 0.628 | 0.970 | 1,251,934 | 3,247 | 0.340 | 0.320 | 0.301 | 0.391 | 0.925 |
| 23 | FOCUS GROUPS | 2,534,711 | 10,268 | 0.782 | 0.463 | 0.696 | 0.892 | 0.992 | 1,251,934 | 523 | 0.167 | 0.500 | 0.094 | 0.744 | 0.990 |
| 24 | FOLLOW UP STUDIES | 2,534,711 | 116,892 | 0.496 | 0.268 | 0.553 | 0.450 | 0.904 | 1,251,934 | 5685 | 0.038 | 0.592 | 0.055 | 0.029 | 0.818 |
| 25 | GENOME WIDE ASSOCIATION STUDY | 2,534,711 | 10,703 | 0.727 | 0.311 | 0.753 | 0.703 | 0.988 | 1,251,934 | 3,054 | 0.533 | 0.311 | 0.537 | 0.530 | 0.982 |
| 26 | HISTORICAL ARTICLE | 2,534,711 | 12,357 | 0.645 | 0.488 | 0.722 | 0.584 | 0.959 | 1,251,934 | 4,067 | 0.610 | 0.488 | 0.642 | 0.581 | 0.972 |
| 27 | HUMAN EXPERIMENTATION | 2,534,711 | 231 | 0.465 | 0.122 | 0.619 | 0.372 | 0.986 | 1,251,934 | 15 | 0.125 | 0.122 | 0.083 | 0.250 | 0.991 |
| 28 | INTERVIEW | 2,534,711 | 435 | 0.689 | 0.333 | 0.747 | 0.639 | 0.992 | 1,251,934 | 122 | 0.602 | 0.333 | 0.657 | 0.556 | 0.976 |
| 29 | INTERVIEWS AS TOPIC | 2,534,711 | 16,920 | 0.486 | 0.268 | 0.460 | 0.514 | 0.979 | 1,251,934 | 1017 | 0.094 | 0.186 | 0.052 | 0.502 | 0.989 |
| 30 | LECTURES | 2,534,711 | 416 | 0.575 | 0.293 | 0.645 | 0.519 | 0.940 | 1,251,934 | 38 | 0.455 | 0.293 | 0.319 | 0.793 | 0.981 |
| 31 | LEGAL CASES | 2,534,711 | 285 | 0.478 | 0.357 | 0.465 | 0.491 | 0.985 | 1,251,934 | 10 | 0.170 | 0.067 | 0.102 | 0.500 | 0.942 |
| 32 | LETTER | 2,534,711 | 5,035 | 0.318 | 0.167 | 0.863 | 0.195 | 0.833 | 1,251,934 | 2,396 | 0.027 | 0.032 | 0.046 | 0.019 | 0.752 |
| 33 | LONGITUDINAL STUDIES | 2,534,711 | 40,491 | 0.666 | 0.301 | 0.701 | 0.635 | 0.952 | 1,251,934 | 2,901 | 0.257 | 0.301 | 0.183 | 0.431 | 0.915 |
| 34 | MATCHED PAIR ANALYSIS | 2,534,711 | 602 | 0.576 | 0.333 | 0.815 | 0.445 | 0.957 | 1,251,934 | 39 | 0.085 | 0.111 | 0.057 | 0.167 | 0.562 |
| 35 | META ANALYSIS | 2,534,711 | 53,043 | 0.877 | 0.394 | 0.885 | 0.870 | 0.991 | 1,251,934 | 3,830 | 0.358 | 0.118 | 0.239 | 0.713 | 0.974 |
| 36 | MULTICENTER STUDY | 2,534,711 | 84,915 | 0.619 | 0.333 | 0.703 | 0.553 | 0.936 | 1,251,934 | 4363 | 0.103 | 0.396 | 0.067 | 0.228 | 0.849 |
| 37 | NEWS | 2,534,711 | 808 | 0.400 | 0.455 | 0.868 | 0.260 | 0.932 | 1,251,934 | 258 | 0.084 | 0.127 | 0.079 | 0.089 | 0.624 |
| 39 | PERSONAL NARRATIVES | 2,534,711 | 492 | 0.312 | 0.875 | 0.805 | 0.193 | 0.973 | 1,251,934 | 34 | 0.067 | 0.261 | 0.049 | 0.105 | 0.968 |
| 40 | PORTRAITS | 2,534,711 | 1,124 | 0.525 | 0.299 | 0.585 | 0.477 | 0.988 | 1,251,934 | 806 | 0.555 | 0.128 | 0.456 | 0.708 | 0.992 |
| 41 | PRACTICE GUIDELINE | 2,534,711 | 3,669 | 0.564 | 0.261 | 0.549 | 0.579 | 0.993 | 1,251,934 | 140 | 0.193 | 0.204 | 0.123 | 0.448 | 0.973 |
| 42 | PREDICTIVE VALUE OF TESTS | 2,534,711 | 47,699 | 0.463 | 0.227 | 0.503 | 0.429 | 0.925 | 1,251,934 | 2834 | 0.065 | 0.182 | 0.043 | 0.136 | 0.892 |
| 43 | PROSPECTIVE STUDIES | 2,534,711 | 139,033 | 0.727 | 0.406 | 0.753 | 0.702 | 0.954 | 1,251,934 | 8,572 | 0.260 | 0.268 | 0.164 | 0.625 | 0.957 |
| 45 | RANDOM ALLOCATION | 2,534,711 | 5,790 | 0.347 | 0.134 | 0.856 | 0.218 | 0.844 | 1,251,934 | 14,736 | 0.195 | 0.040 | 0.143 | 0.305 | 0.839 |
| 46 | RANDOMIZED CONTROLLED TRIAL | 2,534,711 | 110,400 | 0.771 | 0.220 | 0.802 | 0.743 | 0.964 | 1,251,934 | 8,335 | 0.245 | 0.362 | 0.180 | 0.383 | 0.942 |
| 47 | REPRODUCIBILITY OF RESULTS | 2,534,711 | 82,353 | 0.575 | 0.277 | 0.633 | 0.527 | 0.932 | 1,251,934 | 24,681 | 0.279 | 0.191 | 0.228 | 0.360 | 0.869 |
| 49 | RETROSPECTIVE STUDIES | 2,534,711 | 248,643 | 0.782 | 0.397 | 0.812 | 0.755 | 0.963 | 1,251,934 | 16,152 | 0.278 | 0.397 | 0.176 | 0.661 | 0.965 |
| 50 | REVIEW | 2,534,711 | 404,549 | 0.846 | 0.462 | 0.831 | 0.862 | 0.975 | 1,251,934 | 64,304 | 0.609 | 0.618 | 0.543 | 0.693 | 0.955 |
| 52 | SYSTEMATIC REVIEW AS TOPIC | 2,534,711 | 1,970 | 0.403 | 0.062 | 0.339 | 0.497 | 0.981 | 1,251,934 | 111 | 0.233 | 0.784 | 0.438 | 0.159 | 0.994 |
| 53 | TWIN STUDY | 2,534,711 | 2,053 | 0.751 | 0.400 | 0.758 | 0.744 | 0.994 | 1,251,934 | 67 | 0.063 | 0.667 | 0.032 | 1.000 | 1.000 |
| 54 | VALIDATION STUDIES | 2,534,711 | 23,182 | 0.590 | 0.328 | 0.613 | 0.568 | 0.968 | 1,251,934 | 0 | 0.259 | 0.118 | 0.199 | 0.373 | 0.908 |
| 55 | SYSTEMATIC REVIEW | 2,534,711 | 60,871 | 0.911 | 0.941 | 0.885 | 0.939 | 0.997 | 1,251,934 | 4,631 | 0.339 | 0.941 | 0.211 | 0.856 | 0.993 |

## Notes

### Competing Interest Statement

The authors have declared no competing interest.

### Author Declarations

This is bibliographic publication research, and therefore not human subject research.

## References

1. Bastian H, Glasziou P, Chalmers I. Seventy-five trials and eleven systematic reviews a day: how will we ever keep up? PLoS Med. 2010 Sep;7(9):e1000326.

2. Harker J, Kleijnen J. What is a rapid review? A methodological exploration of rapid reviews in Health Technology Assessments. Int J Evid Based Healthc. 2012 Dec;10(4):397–410.

3. Khangura S, Konnyu K, Cushman R, Grimshaw J, Moher D. Evidence summaries: the evolution of a rapid review approach. Syst Rev. 2012;1:10.

4. Rathbone J, Hoffmann T, Glasziou P. Faster title and abstract screening? Evaluating Abstrackr, a semi-automated online screening program for systematic reviewers. Syst Rev. 2015 Jun 15;4(1):1–7.

5. Cohen AM, Hersh WR, Peterson K, Yen PY. Reducing workload in systematic review preparation using automated citation classification. J Am Med Inf Assoc. 2006;13(2):206– 19.

6. Wallace BC, Dahabreh IJ, Schmid CH, Lau J, Trikalinos TA. Modernizing the systematic review process to inform comparative effectiveness: tools and methods. J Comp Eff Res. 2013 May;2(3):273–82.

7. Bae J-M, Kim EH. Citation Discovery Tools for Conducting Adaptive Meta-analyses to Update Systematic Reviews. J Prev Med Pub Health. 2016 Mar 14;49(2):129–33.

8. Cohen AM, Adams CE, Davis JM, Yu C, Yu PS, Meng W, et al. Evidence-based medicine, the essential role of systematic reviews, and the need for automated text mining tools. In: Proceedings of the ACM international conference on Health informatics - IHI ‘10 [Internet]. Arlington, Virginia, USA; 2010. p. 376. Available from: http://portal.acm.org/citation.cfm?doid=1882992.1883046

9. Hartling L, Bond K, Harvey K, Santaguida PL, Viswanathan M, Dryden DM. Developing and Testing a Tool for the Classification of Study Designs in Systematic Reviews of Interventions and Exposures. Agency Healthc Res Qual Dec 2010 Methods Res Rep AHRQ Publ No 11-EHC-007. 2010;

10. Lowe HJ, Barnett GO. Understanding and using the medical subject headings (MeSH) vocabulary to perform literature searches. JAMA. 1994 Apr 13;271(14):1103–8.

11. Aronson AR, Mork JG, Gay CW, Humphrey SM, Rogers WJ. The NLM indexing initiative’s medical text indexer. Medinfo. 2004;89.

12. Huang M, Neveol A, Lu Z. Recommending MeSH terms for annotating biomedical articles. J Am Med Inform Assoc JAMIA. 2011;18(5):660–7.

13. Liu K, Peng S, Wu J, Zhai C, Mamitsuka H, Zhu S. MeSHLabeler: improving the accuracy of large-scale MeSH indexing by integrating diverse evidence. Bioinformatics. 2015 Jun 15;31(12):i339–47.

14. Peng S, You R, Wang H, Zhai C, Mamitsuka H, Zhu S. DeepMeSH: deep semantic representation for improving large-scale MeSH indexing. Bioinforma Oxf Engl. 2016 15;32(12):i70–9.

15. Peng S, Mamitsuka H, Zhu S. MeSHLabeler and DeepMeSH: Recent Progress in Large-Scale MeSH Indexing. In: Mamitsuka H, editor. Data Mining for Systems Biology: Methods and Protocols [Internet]. New York, NY: Springer; 2018 [cited 2021 Feb 9]. p. 203–9. (Methods in Molecular Biology). Available from: https://doi.org/10.1007/978-1-4939-8561-6_15

16. Dai S, You R, Lu Z, Huang X, Mamitsuka H, Zhu S. FullMeSH: improving large-scale MeSH indexing with full text. Bioinformatics. 2020;36(5):1533–41.

17. Xun G, Jha K, Yuan Y, Wang Y, Zhang A. MeSHProbeNet: a self-attentive probe net for MeSH indexing. Bioinforma Oxf Engl. 2019 Oct 1;35(19):3794–802.

18. Marshall IJ, Wallace BC. Toward systematic review automation: a practical guide to using machine learning tools in research synthesis. Syst Rev. 2019 Jul 11;8(1):163.

19. Cohen AM, Smalheiser NR, McDonagh MS, Yu C, Adams CE, Davis JM, et al. Automated confidence ranked classification of randomized controlled trial articles: an aid to evidence-based medicine. J Am Med Inform Assoc JAMIA. 2015 May;22(3):707–17.

20. Wallace BC, Noel-Storr A, Marshall IJ, Cohen AM, Smalheiser NR, Thomas J. Identifying reports of randomized controlled trials (RCTs) via a hybrid machine learning and crowdsourcing approach. J Am Med Inform Assoc JAMIA. 2017 May 25;

21. Przybyła P, Brockmeier AJ, Kontonatsios G, Le Pogam M-A, McNaught J, von Elm E, et al. Prioritising references for systematic reviews with RobotAnalyst: a user study. Res Synth Methods. 2018;9(3):470–88.

22. Wallace BC, Small K, Brodley CE, Lau J, Trikalinos TA. Deploying an interactive machine learning system in an evidence-based practice center: abstrackr. In: Proceedings of the 2nd ACM SIGHIT International Health Informatics Symposium. ACM; 2012. p. 819–24.

23. Shemilt I, Khan N, Park S, Thomas J. Use of cost-effectiveness analysis to compare the efficiency of study identification methods in systematic reviews. Syst Rev. 2016;5(1):1–13.

24. Wieland LS, Robinson KA, Dickersin K. Understanding why evidence from randomised clinical trials may not be retrieved from Medline: comparison of indexed and non-indexed records. BMJ. 2012;344:d7501.

25. Glanville JM, Lefebvre C, Miles JNV, Camosso-Stefinovic J. How to identify randomized controlled trials in MEDLINE: ten years on. J Med Libr Assoc JMLA. 2006 Apr;94(2):130– 6.

26. Salvador-Oliván JA, Marco-Cuenca G, Arquero-Avilés R. Errors in search strategies used in systematic reviews and their effects on information retrieval. J Med Libr Assoc JMLA. 2019 Apr;107(2):210–21.

27. Bekhuis T, Demner-Fushman D, Crowley RS. Comparative effectiveness research designs: an analysis of terms and coverage in Medical Subject Headings (MeSH) and Emtree. J Med Libr Assoc JMLA. 2013 Apr;101(2):92–100.

28. Cohen AM, Dunivin ZO, Smalheiser NR. A probabilistic automated tagger to identify human-related publications. Database J Biol Databases Curation. 2018 01;2018:1–8.

29. Marshall I, Storr AN, Kuiper J, Thomas J, Wallace BC. Machine Learning for Identifying Randomized Controlled Trials: an evaluation and practitioner’s guide. Res Synth Methods. 2018;9.4:602–14.

30. Search Strategy Used to Create the PubMed Systematic Reviews Filter [Internet]. U.S. National Library of Medicine; [cited 2020 Oct 14]. Available from: https://www.nlm.nih.gov/bsd/pubmed_subsets/sysreviews_strategy.html

31. Smalheiser NR, Cohen AM. Design of a generic, open platform for machine learning-assisted indexing and clustering of articles in PubMed, a biomedical bibliographic database. Data Inf Manag. 2018;2(1):27–36.

32. Smalheiser NR, Cohen AM, Bonifield G. Unsupervised Low-Dimensional Vector Representations for Words, Phrases and Text that are Transparent, Scalable, and produce Similarity Metrics that are not Redundant with Neural Embeddings. J Biomed Inform. 2019 Jan 14;103096.

33. Hashimoto K, Kontonatsios G, Miwa M, Ananiadou S. Topic detection using paragraph vectors to support active learning in systematic reviews. J Biomed Inform. 2016 Aug 1;62:59–65.

34. D′Souza JL, Smalheiser NR. Three Journal Similarity Metrics and Their Application to Biomedical Journals. PLOS ONE. 2014 Dec 23;9(12):e115681.

35. Chang C-C, Lin C-J. LIBSVMLJ: a library for support vector machines [Internet]. 2006. Available from: Software available at http://www.csie.ntu.edu.tw/~cjlin/libsvm

36. Smalheiser NR, Fragnito DP, Tirk EE. Anne O’Tate: Value-added PubMed search engine for analysis and text mining. PloS One. 2021;16(3):e0248335.

37. Schneider J, Hoang L, Kansara Y, Cohen AM, Smalheiser NR. Evaluation of publication type tagging as a strategy to screen randomized controlled trial articles in preparing systematic reviews. JAMIA Open. (in press).

38. Cohen AM, Smalheiser NR. UIC/OHSU CLEF 2018 Task 2 Diagnostic Test Accuracy Ranking using Publication Type Cluster Similarity Measures. In: CLEF 2018 Working Notes: Working Notes of CLEF 2018 - Conference and Labs of the Evaluation Forum, Editors Linda Cappellato, Nicola Ferro, Jian-Yun Nie, Laure Soulier. Avignon, FRANCE;

39. Marshall IJ, Kuiper J, Wallace BC. RobotReviewer: evaluation of a system for automatically assessing bias in clinical trials. J Am Med Inform Assoc. 2016;23(1):193–201.

